# Challenges in Estimating Time-Varying Epidemic Severity Rates from Aggregate Data

**DOI:** 10.1101/2024.12.27.24319518

**Authors:** Jeremy Goldwasser, Addison J. Hu, Alyssa Bilinski, Daniel J. McDonald, Ryan J. Tibshirani

**Affiliations:** Department of Statistics, University of California, Berkeley; Department of Statistics, Carnegie Mellon University; Departments of Health Policy and Biostatistics, Brown University; Department of Statistics, University of British Columbia

**Keywords:** Severity rate, case-fatality rate, epidemiology, statistical bias, COVID-19, aggregate data

## Abstract

Severity rates like the case-fatality rate and hospitalization-fatality rate are key metrics in public health. To guide decision-making in response to changes like new variants or vaccines, it is imperative to understand how these rates shift in real time. In practice, time-varying severity rates are typically estimated using a ratio of aggregate counts. We demonstrate that these estimators are capable of exhibiting large statistical biases, with concerning implications for public health practice, as they may fail to detect heightened risks or falsely signal nonexistent surges. We supplement our mathematical analyses with experimental results on real and simulated COVID-19 data. Finally, we discuss strategies to mitigate this bias, drawing connections with effective reproduction number (*R_t_*) estimation.

## 1 Introduction

Several public health metrics of interest express the probability that a second, often more severe outcome will follow a primary event. We refer to such metrics as “severity rates.” Important examples of severity rates are the hospitalization-fatality rate (HFR) and case-fatality rate (CFR), commonly used to assess the deadliness of an epidemic^(1,2,3,4)^.

In an ideal setting, severity rates can be obtained directly from a line-list or claims data set containing individual patient outcomes. Such line-lists do not necessarily need to be comprehensive, or even representative of the target population of interest. Various techniques can be used to adjust for biases in non-representative line-lists, such as post-stratification on characteristics like age ^(5)^, race ^(6)^, and seroprevalence ^(3)^. However, real-time tracking of individuals in fast-moving epidemics has in many settings been infeasible, even at smaller scales. For example, the CDC’s line-list during the COVID-19 pandemic excluded large geographic regions, was only updated monthly, and had missing death statuses for a high proportion of individuals ^(7)^. Techniques like importance reweighting are often insufficient to address these problems.

When real-time severity rates cannot be discerned from line-lists, they are routinely estimated from aggregate count data. Aggregate case and death counts were widely used to estimate and report COVID-19 CFRs in the academic literature ^(8,9,10,11)^, as well as in major news outlets including The Atlantic ^(12)^, Wall Street Journal ^(13)^, and New York Times^(14)^. They were also used by public health organizations like the CDC^(15,16)^, and reported by the Trump and Biden administrations ^(17,18)^.

Several works assume severity rates are constant in time ^(19,20,21,22,23)^. However, consequential shifts can occur in response to factors such as new therapeutics, vaccines, and variants. Therefore, many works estimate time-varying severity rates from aggregate data streams. Standard real-time approaches do so with a ratio of primary and secondary counts. In fact, ratio estimators are so common that CFR is often (mis)labeled the case-fatality *ratio* ^(2)^.

In this work, we show that these ratio estimators are prone to nontrivial statistical bias. We empirically validate our mathematical analysis, tracking the hospitalization-fatality rate (HFR) during COVID-19. Bias arises as a consequence of changing severity rates— precisely what time-varying estimates are employed to detect. For example, the ratio estimators failed to quickly signal increased risk in the onset of the Delta wave. Bias also arises due to misspecification of the delay distribution that relates primary and secondary events. This is particularly troublesome during various stages of a surge, especially for the popular lagged ratio. Once again, these are periods in which it is critical to properly ascertain the severity rate. While the HFRs dropped in the aftermath of the initial Omicron surge, the lagged ratio’s estimates jumped 50% above the true values. These experiments use hospitalization data, but our analysis applies broadly to the bias of all severity rates. Indeed, we rerun our simulation experiments on CFR, delivering even more biased results.

We provide practical heuristics for when epidemiologists should expect this bias in practice. These may prevent poor decision-making based on biased estimates of severity rates. More broadly, our work exposes the limitations of existing approaches, motivating the use of alternatives. We point towards methodology which may avoid this bias, and offer suggestions to improve it. Code to reproduce our experimental results is available at https://github.com/jeremy-goldwasser/Severity-Bias.

## 2 Methods

In this section, we introduce the main estimators we study, and analyze their bias. Subsequently, we detail the data used for empirical study and validation.

### 2.1 Severity rate estimators

Severity rates convey the probability that a primary event will result in a secondary event in the future. In the case of CFR, for example, a primary event is a positive COVID-19 case and a secondary event is a death with a positive test result. Formally, a time-varying severity rate at time *t* is generally defined as:

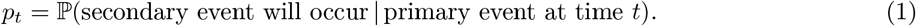

Here, *t* may represent a discrete interval of time, such as a given day or week. It also may be understood in a continuous-time fashion. Although this will not be our focus in this paper, the same general principles apply in the continuous-time case. For simplicity, we will consider only the discrete-time setting, and we index time steps via integers, as in *t* = 0, 1, 2, … .

Throughout, we denote by {*X*_*t*_} and {*Y*_*t*_} the aggregate time series of new primary and secondary events, respectively. These are often counts, and we will generally refer to them as such. At time *t*, we assume data for all past *s* ≤ *t* is available, but future data is not. Therefore real-time estimates of *p*_*t*_ can only rely on past counts {*X*_*s*_} _*s*≤*t*_ and {*Y*_*s*_} _*s*≤*t*_. In practice, to stabilize estimates, smoothed counts are often used in place of raw counts. This may be simply absorbed into the notation for *X*_*t*_ and *Y*_*t*_, and we do not address smoothing explicitly in the formulation of the estimators, but refer back to this issue in Section 2.4.

In the context of CFR, for example, {*X*_*t*_} and {*Y*_*t*_} denote cases and deaths. However, the following estimators and their biases are shared for any severity rate *p*_*t*_, including CFR and HFR.

### Lagged estimator

The canonical estimator for time-varying severity rates is a ratio between the counts of primary and secondary events, offset by a lag 𝓁. This estimator is widely-used in epidemiology, both in the academic literature and in public health practice and communication (e.g., ^13,12,8,2,24,9,10,11^). For concreteness, we define the *lagged ratio* at time *t* as:

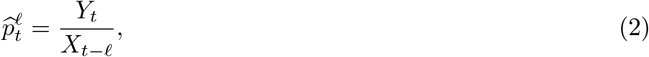

where 𝓁 ≥ 0 is a given parameter (often chosen to maximize cross-correlation between {*X*_*t*_} and {*Y*_*t*_}). Note without loss of generality that the lag itself may be time-varying, adapting to shifts in the delay distribution.

### Convolutional estimator

Alternative methods for estimating severity rates utilize a delay distribution, which relates the two time series. The delay distribution at time *t* and lag *k* is defined as:

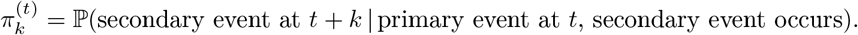

Throughout this work, we assume that the delay distribution has a finite support of *d* time steps. For the sake of notational convenience, we also assume the delay distribution is stationary: 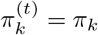 for all *k* and *t*. However, our analyses hold in the case of time-varying delay distributions. While the delay distribution is generally unknown, several tools exist to estimate them from aggregate or line-list data; see ^25^ for a review. Given *π*, we can express the expected number of secondary events at time *t* as follows:

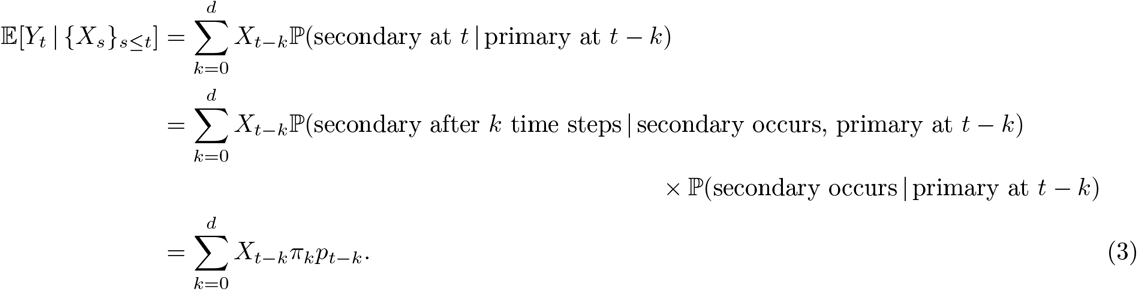

This is a convolution of the delay distribution against the product of primary incidence and the severity rate. If the severity rate remains constant, *p*_*t*−*k*_ = *p*_*t*_ for all *k* between 0 and *d*, then the expression in (3) simplifies to 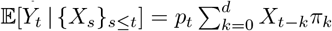. As studied in Overton et al. ^(26)^, we can rearrange this relationship in order to estimate the severity rate at *t*, after plugging-in an estimate *γ* of the delay distribution *π*:

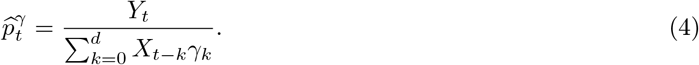

We call this the *convolutional ratio* estimator of the severity rate. The superscript in our notation 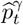 emphasizes that *γ* is the distribution used in the definition of the estimator (4). To reiterate, in moving from (3) to (4), we are implicitly assuming that the severity rate *p*_*t*_ is stationary over the interval of time from *t − d* and *t*. Of course, this runs in contradiction to the fact that we are trying to estimate a time-varying severity rate in the first place. As we will see shortly, this can create significant bias in the convolutional ratio.

Some further comments are in order. The convolutional ratio has a longer history of study and use in the literature on estimating stationary severity rates. Indeed, Nishiura et al. ^(27)^ developed the estimator in this setting, and used it to analyze the CFR in the H1N1 influenza pandemic of 2009. (The main difference to (4) is that in the stationary case we aggregate both the numerator and denominator over all past data.) This estimator, which is sometimes called the *delay-adjusted* estimator of the severity rate, is popular in the academic literature and public health practice (e.g., ^19,5,28,29^), though arguably less popular than the lagged ratio. The package `cfr`^(30)^ gives an R implementation, for both the stationary and time-varying cases.

Furthermore, we note that (4) can be seen as a generalization of the lagged ratio estimator (2): when we take *γ* to be a point mass at lag 𝓁, i.e., *γ*_𝓁_ = 1 and *γ*_*k*_ = 0 for all *k* ≠ 𝓁, then (4) reduces to (2).

### Connection with reproduction numbers

Severity rates bear a natural connection with reproduction numbers. Both the true severity rate as defined in (1) and the case reproduction number *R*_*t*_ are defined as the average number of secondary events produced by a single primary event at *t*, but in contrast to severity rates, reproduction numbers have infections as the primary and secondary events, where a single infection can produce more than one follow-on infection. Playing the role of the delay distribution *π* in our setting is the generation interval distribution in reproduction numbers, which measures the time in between primary and secondary infections.

Severity rates and reproduction numbers can also be estimated similarly. Because primary events at *t* produce secondary events after *t*, their effect is not observed in real time. Therefore, standard real-time estimators for severity rates (2), (4) and for *R*_*t*_ both analyze the number of secondary events at *t* produced by relevant primary events. In words, this adopts a “backward-looking” perspective that is possible in real time, rather the the “forward-looking” perspective inherent to the definition in (1).

For reproduction numbers, the backward-looking quantity has its own name: *instantaneous R*_*t*_, which is the average number of secondary infections at time *t* produced by a single primary infection in the past. One of the most popular traditional estimators of instantaneous *R*_*t*_ is based on an essentially identical idea to the convolutional ratio ^(31,32)^:

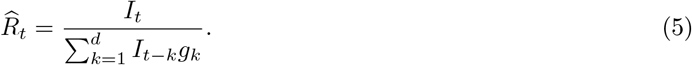

Here, *I*_*t*_ denotes the number of new infections at *t*, and *g* is the generation interval distribution. Note that the difference between (4) and (5) is that the latter uses the same aggregate time series, infections, in both the numerator and denominator. While more modern frameworks for estimating instantaneous *R*_*t*_ are often Bayesian (e.g., ^33^), the underlying basic point estimates are still the same.

### 2.2 Well-specified analysis

First we analyze the bias of the convolutional ratio (4) in what we call the *well-specified* case, where the true delay distribution *π* is known. Formally, for an estimator 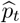 of *p*_*t*_, we define its bias as:

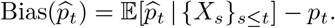

#### Proposition 1

*Assume the true delay distribution π is known. The bias of the well-specified convolutional ratio* 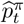 *is*

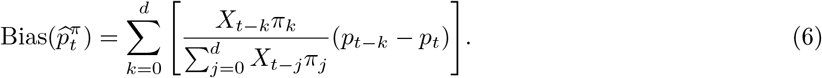

The proof of Proposition 1 is given in Section S1.1 of the Supplement. Intuitively, the well-specified bias can be interpreted as a weighted average of the difference between trailing and current severity rates. The weights depend on the delay distribution and primary incidence curve. Figure 1 provides an accompanying illustration.

**Figure 1:**
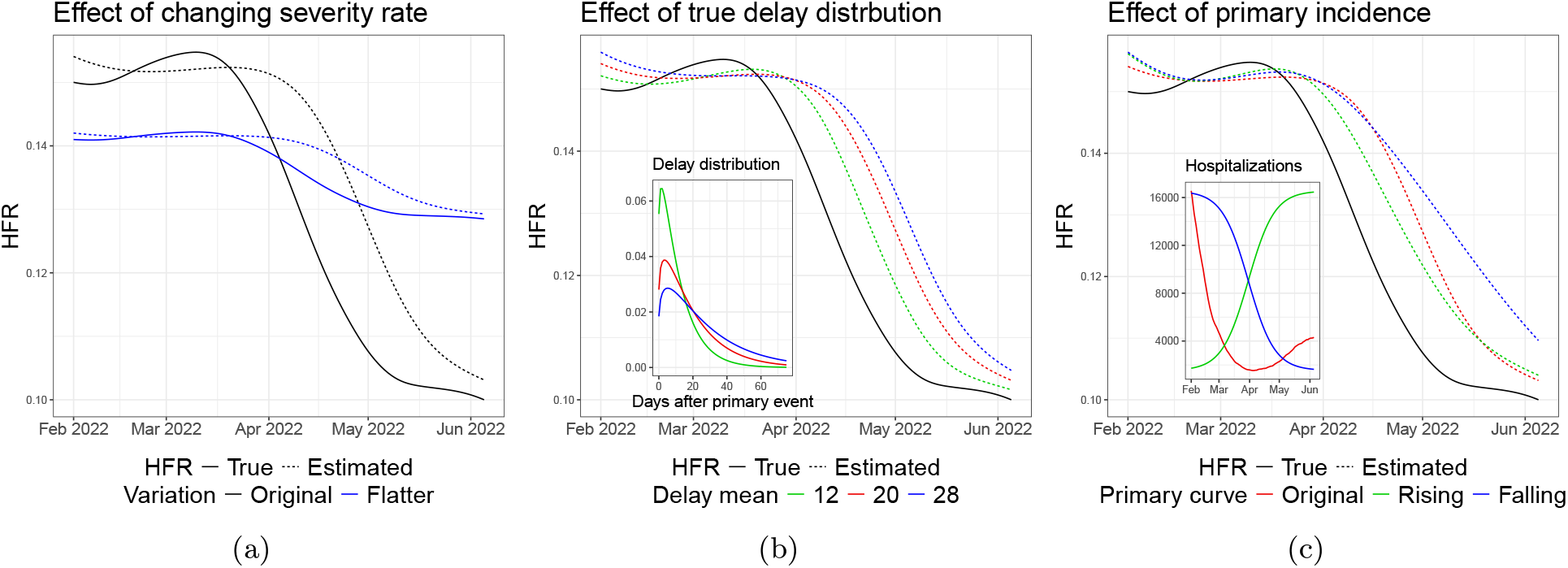
Simple examples which illustrate the effects of the three factors explained above on the bias of the well-specified convolutional ratio (6). In each figure, the colors correspond to different versions of the factor of interest. The primary incidence curve measures COVID-19 hospital admissions, as reported to the HHS in early 2022. We then simulate a secondary incidence curve, COVID-19 deaths, from (3), without noise. The underlying HFR curve *p*_*t*_ and delay distribution *π* used (in simulating deaths) were derived from external data sources, as explained in more detail in Section 2.4.

1. **Changes in severity rate**. The central component of this bias expression is the difference *p*_*t*−*k*_ − *p*_*t*_. When the severity rate is constant over the *d* preceding time points, the convolutional ratio is unbiased (because this difference is zero). This falls in line with the motivation used to derive this estimator, as explained in the last subsection. But when severity rates change before *t*, these difference terms will be nonzero, in which case the estimator will be generally biased. Figure 1a shows a simple example of this: the estimated severity rates are most inaccurate in periods where the true rate is changing quickly. To make matters worse, the bias is in the opposite direction of the trend we want to detect; for example, suppose the severity rate is monotonically falling, with *p*_*t*_ *< p*_*t*−1_ *<* … < *p*_*t*−*d*_. The bias will then be positive, meaning the ratio estimates do not decline with the true rate. In fact, the estimated severity may even rise, not fall. Conversely, when true severity rates are rising, the estimates will be too low.
2. **The delay distribution**. How much the changing severity rates impact the bias depends on the shape of the delay distribution *π*. In general, the bias will be greatest when the delay distribution has a long enough tail to upweight significant differences in severity rate. While this distinction may appear subtle, Section 3 highlights its surprisingly large effects. The simple example in Figure 1b also shows significant differences in bias between shorter and longer delay distributions.
3. **The primary incidence curve**. Changing primary incidence *X*_*t*_ will also affect the bias, presuming the severity rate changes roughly monotonically in the recent past. Intuitively, this up- or down-weights the terms *X*_*t*−*k*_*π*_*k*_(*p*_*t*−*k*_ − *p*_*t*_) at times further from the present, which are likely to contribute the most bias. In general, falling primary incidences will amplify the bias, whereas rising events will minimize it. Figure 1c provides an illustration of this phenomenon.

Section S2.1 of the Supplement provides further analysis, by discussing simplified settings in which the bias (6) described in Proposition 1 itself simplifies in elucidating ways.

### 2.3 Misspecified analysis

We now analyze the bias of the convolutional ratio (4) for an arbitrary distribution *γ*. Recall that *π* denotes the true delay distribution in (3). We refer to the present as the *misspecified* case, as *γ* may differ from *π*.

#### Proposition 2.

*The bias of the convolutional ratio* 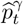 *(where the true delay distribution π is unknown, and the working delay distribution γ is arbitrary, but also supported on d time steps) is*

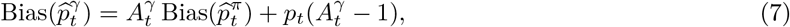

*where* 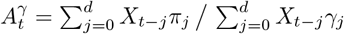. *This compares how the delay distributions convolve against the most recent primary incidence levels*.

The proof of Proposition 2 is given in Section S1.2 of the Supplement. Before delving into the technical details, we first summarize its implications for the lagged ratio. When primary incidence 𝓁 days ago is less than the average count weighted by the delay distribution, the lagged ratio is typically above the well-specified convolutional ratio. Otherwise, when counts 𝓁 days ago exceed the delay-weighted average, it tends to lie below. This leads to the following heuristics, which we justify subsequently.

- **Heuristic #1:** When primary incidence is climbing rapidly, the lagged ratio tends to overestimate severity, at least relative to the well-specified convolutional ratio.
- **Heuristic #2:** As incidence falls swiftly, the lagged ratio may drop just as sharply, even if true severity has not changed.
- **Heuristic #3:** Once incidence stabilizes after a decline, the lagged ratio can exhibit sudden, counter-intuitive jumps.

Under misspecification, the proposition gives an additive decomposition (7) of the convolutional ratio bias, based on the well-specified bias Bias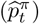 (as studied in Proposition 1), and a misspecification factor 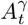 . At the outset, we note that if *π* = *γ* (no misspecification), we have 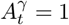, and (7) reduces to the well-specified bias. Generally, values of 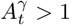 amplify the oracle bias and add positive misspecification bias 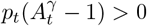; meanwhile, values of 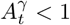 shrink the oracle bias and add negative misspecification bias 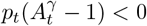 .

Whether or not misspecification contributes a larger magnitude of bias overall hence depends on whether or not the sign of the misspecification term 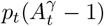 agrees with the sign of the oracle bias Bias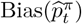. This need not always be the case, though in our experience, it is often true in both real and simulated experiments, as we will see in Section 3. Here, to gain more insight, we study the behavior of the bias in three settings:

- Smooth *γ* with a lighter tail and smaller mean than *π* (more mass concentrated at recent time points).
- Smooth *γ* with a heavier tail and larger mean than *π* (less mass concentrated at recent time points).
- Nonsmooth *γ*, with a point mass at lag 𝓁; we reiterate that in this case the convolutional ratio reduces to the lagged ratio 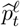 in (2). We also note that its misspecification factor 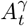 reduces to a quantity we similarly denote 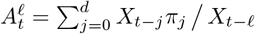, and its bias (7) reduces to:

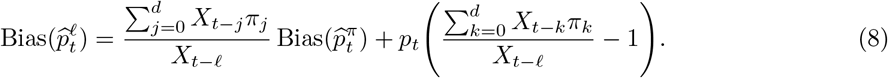

We discuss the behavior of the bias in these three settings, as a function of the primary incidence curve (which drives bias through the misspecification factor 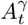). Figure 2 provides an accompanying illustration.

**Figure 2:**
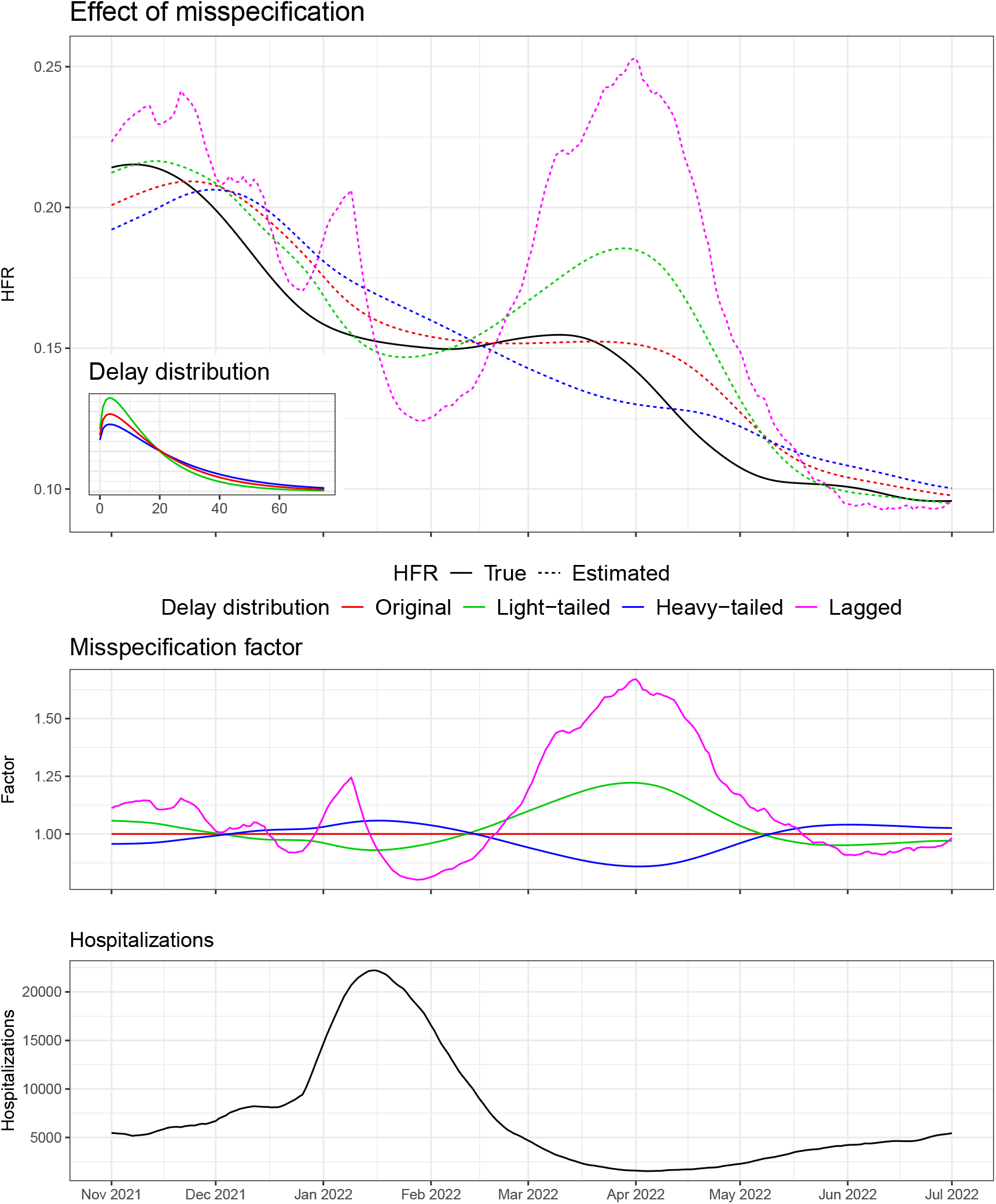
Examples of convolutional ratio estimates under misspecification of the delay distribution. As in Figure 1, the primary events are COVID-19 hospitalizations, as reported to the HHS, and secondary events are deaths simulated noiselessly from (3). The underlying HFR curve *p*_*t*_ and delay distribution *π* used in the simulation were fit using external data sources detailed shortly in Section 2.4. The lagged ratio estimator used 𝓁 = 16, chosen to maximize cross-correlation between hospitalizations and deaths.

#### Primary incidence rising

Consider the case where primary events are rising—first slowly, then rapidly before leveling off. A lighter-tailed *γ* will place more weight on recent time points, with higher counts, than *π*; thus 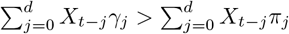, so 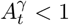. The opposite occurs for a heavier-tailed *γ*: this places more weight on distant low-count time points and less on the ongoing surge, so 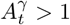. A point mass distribution *γ* places all of its mass on *X*_*t*−𝓁_, and during the steepest phase of the rise, this is considerably less than *X*_*t*_. The true delay *π* distributes mass across the *d* most recent time points, and a large fraction of its mass will be convolved against the last 𝓁 −1 time points, whose counts exceed *X*_*t*−𝓁_. The times before *t* −𝓁 have less of an offsetting effect, because incidence has risen at a growing rate. Hence, during a surge we will see 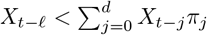, and 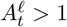. However, the behavior of 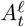 will be generally more erratic than 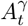 for a smooth distribution *γ*, as the denominator in the former is less smooth as *t* varies.

Figure 2 visualizes this as hopitalizations (primary events) rise between December 2021 and mid-January 2022. Throughout this period 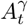 is below/above 1 for the light-tailed/heavy-tailed *γ*. Meanwhile, 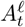 spikes to 1.25 in early January. Correspondly, the lagged HFR rises to 20% when the true one drops to 15%.

#### Primary incidence falling

Next assume primary incidence reaches a maximum and begins to fall. The smooth distributions behave much the same as when incidence was rising. The light-tailed distribution has more mass around the peak than *π*, so 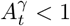. Conversely, 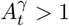 for the heavier-tail distribution because it convolves more mass before the top of the rise. The lagged bias changes its behavior in this period; while 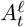 had exceeded 1 before the peak, it quickly plunges below 1. At exactly 𝓁 time points after the peak, the lagged estimator attains the smallest possible value of 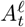, as *X*_*t*−𝓁_ maximizes its denominator. Again, the lagged ratio is likely to have larger fluctuations of 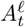, since its denominator reaches extremes that are not witnessed in 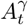 (the convolution in the denominator of 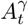 acts as a smoother).

In Figure 2, we can see 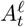 drop below 0.8 near the start of February 2022; this happens precisely 𝓁 = 16 days after daily new hospitalizations peak above 20,000 in mid-January. The lagged ratio falls from 20% to 12.5% accordingly, with the true HFR remains roughly constant, hovering around 15%. In the same period, the convolutional ratios (with light- or heavy-tailed *γ*) stay quite close to the true HFR.

#### Primary incidence levels out from a fall

The most jarring instance of misspecification bias occurs as primary incidence levels out. The true delay distribution *π* has a heavier tail than the low-mean, light-tailed distribution *γ*. It also has a heavier tail than the point mass distribution, which has no tail at all. This has important implications as the peak of the surge fades into the past. Compared to *π*, the light-tailed *γ* and point mass distribution convolve little to no mass with the high-count period of the wave. As a result, both 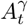 and 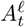 rise above 1, and severity rate estimates spike. The magnitude of this spike depends how quickly primary incidence is changing.

Figure 2 displays this false spike. Around the start of April 2022, we see 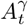 (for light-tailed *γ*) and 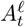 reach maximums near 1.25 and 1.68, respectively. Their corresponding HFR estimates reach 18% and 25% while the true HFR has fallen below 14%. This is of course highly problematic as it signals a rise in severity at a very counterintuitive time, when hospitalizations are at their lowest. The heavy-tailed delay *γ* has the opposite trend and underestimates the true HFR at this time, but by a smaller amount.

### 2.4 Experimental setup

Here we describe the data and general experimental setup used in Figures 1 and 2, and in Section 3.

#### Hospitalization-fatality rate

Our experiments primarily analyze the HFR throughout the COVID-19 pandemic. While HFR may be less common as an object of study compared to CFR, it has a few advantages. Firstly, we were able to find a good “ground truth proxy” for the national HFR during the COVID-19 pandemic, as published by the National Hospital Care Survey (NHCS). This helps guide our simulations and also serves as validation data for us, as we describe in more detail below.

A second advantage is that hospitalization reporting was much more complete than case reporting throughout the pandemic. Hospitals were mandated to report new daily COVID-19 admissions to the Department of Health and Human Services (HHS) ^(34)^. Due to changes in case ascertainment over time (cases as a fraction of infections), it is harder to interpret the CFR in a time-varying fashion, i.e., harder to understand what precisely this is reflecting over the course of the pandemic.

Thirdly, hospitalization counts published by the HHS are aligned by admission date. This makes it more meaningful to interpret the HFR as a reflection of severity, especially in a time-varying fashion. In comparison, case counts as aggregated by John Hopkins University (JHU) ^(35)^ (the central resource for comprehensive COVID-19 case data in the US) are aligned by report date. Extreme reporting delays (sometimes cases were reported 45 days after infections, see, e.g., ^36^) make the CFR less meaningful to study as a time-varying quantity, even outside of ascertainment issues.

To assess the generality of our results, Section S4 of the Supplement conducts the simulated analyses using CFR in place of HFR. In addition to Figure 4 (forthcoming in the results section), Figures 1 and 2 are reproduced using case data. In all instances, the resulting trends mirror those for HFR, confirming that the bias mechanisms identified here apply broadly across severity metrics.

#### Aggregate data streams

To estimate the real-time HFR, we use aggregate counts of daily COVID-19 hospitalizations and deaths as made available in the Epidata API ^(37)^, developed by the Delphi Group. Like HHS for hospitalizations, the JHU Center for Systems Science and Engineering (CSSE) provided the definitive resource for real-time death counts during the pandemic. These counts reflect times at which deaths were reported to health authorities, not necessarily when they actually happened. Hence raw JHU death counts are highly volatile due to reporting idiosyncrasies like day-of-week effects and data dumps. Hospitalizations are also subject to strong day-of-week effects. We thus smooth all data with a 7-day trailing average, for both hospitalizations and deaths.

Our real-time estimates of HFR actually use data that was available two days after the date in question. This was done to account for a typical two-day latency in the most recent data available. In this sense, one can actually view our real-time estimates as a two-day backcast of the HFR. In the rare event that counts were still unavailable at a two-day lag, we imputed their values with the most recently observed data (this is a common scheme, called last-observation-carried-forward or LOCF).

#### Hyperparameters

The ratio estimators of the HFR require choices of the lag 𝓁 and delay distribution *γ*. The experiments in Section 3 use a lag of 𝓁 = 20 days, which roughly maximizes the cross-correlation between hospitalizations and deaths over the entire pandemic. For *γ*, we use a discrete gamma distribution, and set its support length to be *d* = 75 days, a conservative choice. For its mean, we use 20 again; this agrees nicely with a UK analysis that finds a median hospitalization-to-death time of 11 days ^(38)^,^1^ and a CDC analysis that finds 63% of COVID-19 deaths are reported in 10 days ^(39)^. We set the standard deviation to 18, because the delay distributions fit by the UK study had standard deviations that were roughly 90% of their means.

Section S3 of the Supplement evaluates the robustness of findings against different hyperparameter values. We show our findings do not change across a wide range of lags (Figure S7) and delay distributions (Figure S8).

#### Validation data

Due to a lack of ground truth, the bias cannot be rigorously evaluated on real data. However, there are sound ways to better approximate the true HFRs from external sources. One way is to use estimates from the National Hospital Care Survey (NHCS) ^(40)^, which records weekly HFR based on a representative subset of 601 hospitals across the US. These estimates end up being consistently biased downwards because they are only based on deaths which occur in the hospital. A CDC analysis ^(15)^ found that roughly 60% of COVID-19 deaths occurred in hospitals in 2022, down from nearly 70% in 2021 and 2022. To account for non-inpatient deaths, we divide the NHCS estimates by these percentages. Lastly, we smooth the resulting HFR estimates with a spline, via the smooth.spline function in R (which chooses the smoothness hyperparameter by minimizing generalized cross-validation error). This results in our proxy for the ground truth HFR curve *p*_*t*_.

This external estimate is a useful benchmark to judge the fidelity of our HFR estimates. Of course, it is not perfect, and is derived from a relatively small subset of hospitals. Results in Section 3 suggest it may be too high in late 2022. Section S3.1 in the Supplement discusses alternative approximations to the ground truth HFR. These alternatives, shown in Figure S4, all exhibit similar qualitative behavior.

## 3 Results

We study the performance of the ratio estimators in greater depth, on real and simulated data. Throughout, we continue to use COVID-19 hospitalizations reported to the HHS as the primary incidence curve.

### 3.1 COVID-19 data

Figure 3 displays real-time HFR estimates from November 2020 to December 2022, which spans the major COVID-19 waves. These estimates were fit to the data described in Section 2.4. The real-time hospitalization and death counts exhibit a fair degree of instability, and recall, these were preprocessed with a 7-day trailing average. Even then, the convolutional (4) and lagged ratio (2) estmators each had wild spikes, so we further smoothed the estimates from both methods with a 7-day trailing average. Section S3 studies the sensitivity of the results to the choice of smoothing window in postprocessing. Figure S6 shows window size has very little qualitative effect on the main shape of the HFR estimates from either method.

**Figure 3:**
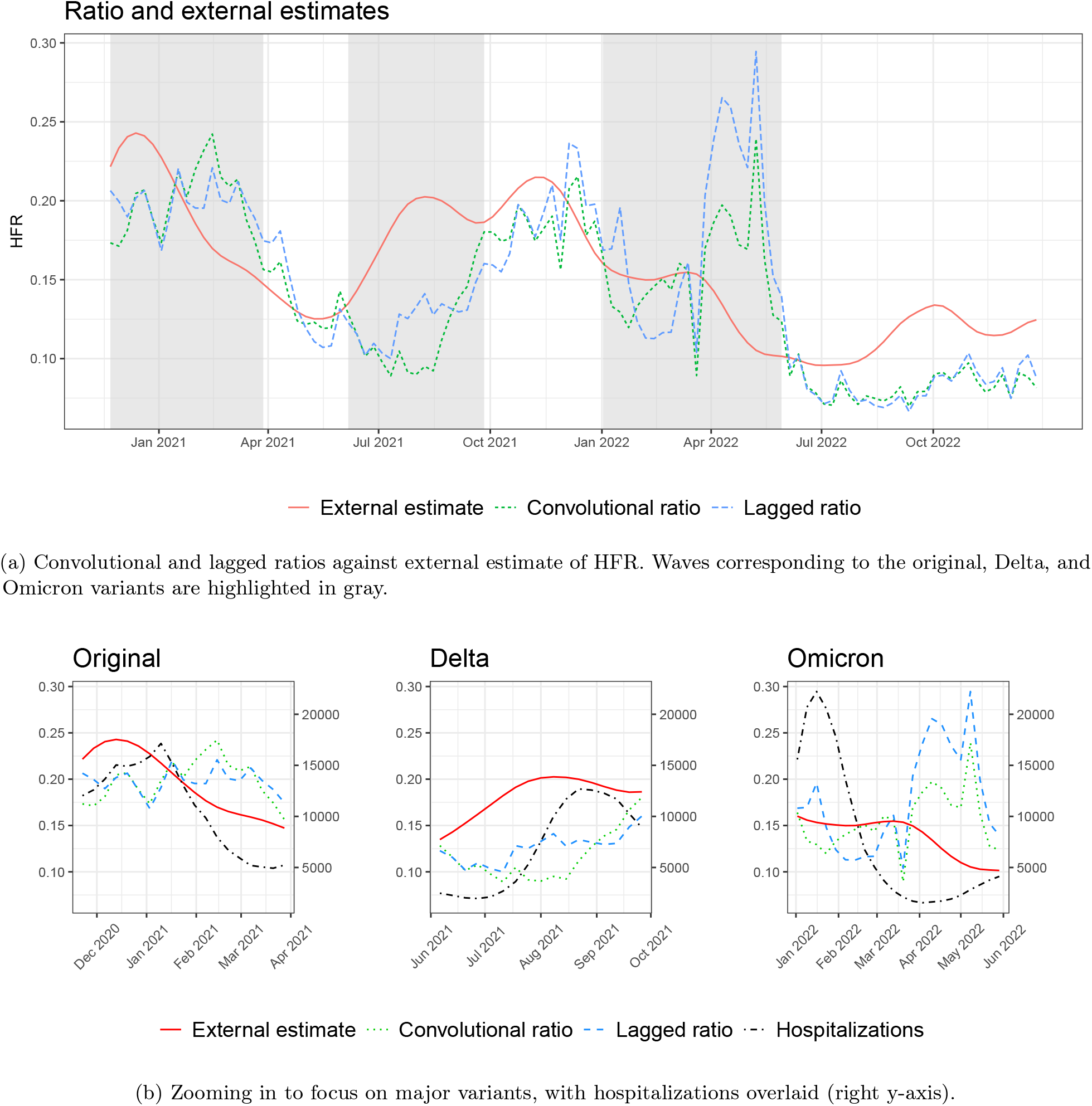
Comparing ratio and external (NHCS-based) estimates of HFR. Computed using real-time COVID-19 counts in the US, from November 2020 through December 2022.

**Figure 4:**
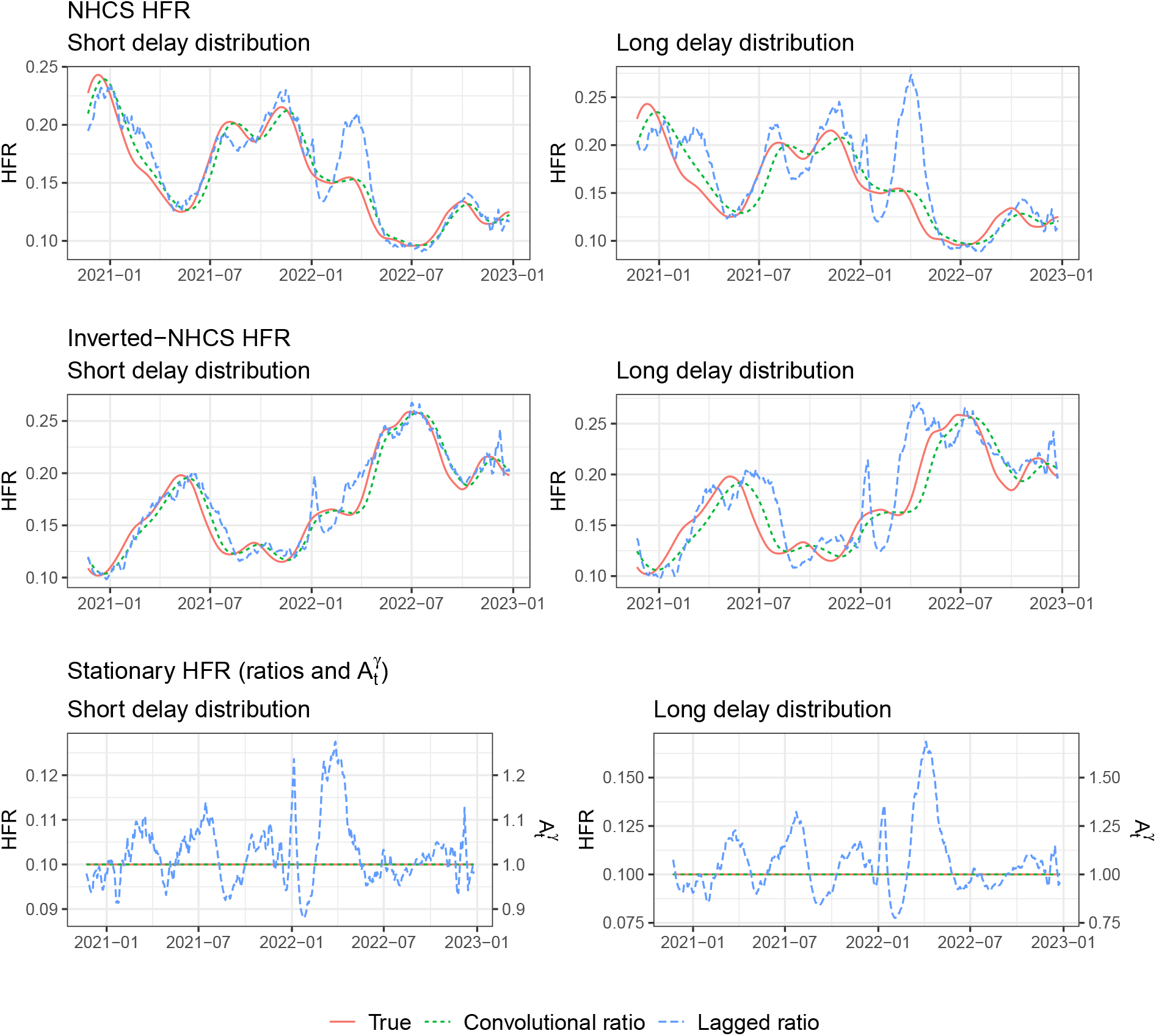
Convolutional and lagged ratios over various simulation settings, with three different underlying HFR curves (rows) and two delays (columns). The last row, for stationary HFR, also provides 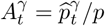.

Overall, both ratio estimators perform poorly—their bias is consistent and nontrivial, especially for the lagged estimator. Both respond very slowly to changes in the HFR. As the HFR declines following the wave in winter 2021, both ratios hover near 20% for several months. More troublingly, they are too slow to detect the rise in HFR in the early Delta period (summer 2021). If the purpose of these estimators is to inform stakeholders of increased risks in real time, they failed during the Delta surge.

The most significant bias comes in the middle of the Omicron wave in spring 2022. In this period, the HFR remains around 15% until April, then sharply declines to 9% two months later. The ratio estimates first fluctuate around the true HFR, and then subsequently, both estimates surge as the true HFR nears its nadir, with the lagged ratio approaching 30%. This dramatic upswing signals a serious false alarm. The analysis in Sections 2.2 and 2.3 explain each of these failure cases.

#### Well-specified analysis

We start by analyzing the convolutional ratio with respect to the well-specified bias expression in Proposition 1. While this expression assumes that the true delay distribution is known, we found that different choices of delay distribution generally yield similar bias; see Figure S8 in Section 3 of the Supplement. This indicates that our convolutional ratio may not be far from the oracle (well-specified) ratio. Proposition 1 indicates that the bias moves in the opposite direction of the true severity rate. This occurs during the Delta wave, when the HFR rises well before the ratio estimates do. On the other hand, falling HFR produces positive bias, as observed in the original and Omicron waves.

The enormity of the bias during Omicron can partially be attributed to the precipitous decline in hospitalizations, as falling primary incidence has been shown to exacerbate the bias. Average daily hospitalizations declined from over 20,000 in mid-January to only 1,500 by April 1. Lastly, the delay distribution is relatively heavy-tailed, because the aggregate deaths here (from JHU) are aligned by report date. We find that this has a substantial impact on the bias, as analyzed in Supplement Section S2.2. Figure S3 shows a large gap in bias between ratios computed on NCHS versus JHU deaths.

#### Misspecified analysis

The misspecification analysis explains central discrepancies between the convolutional and lagged ratios. Section 2.3 discusses why we expect 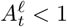 around the start of a decline in primary incidence (Heuristic #2). As a result, the lagged ratio will incur negative misspecification bias, so it takes lower values than the convolutional ratio. We observe this when hospitalizations with the original variant decline from their peak in mid-January 2021. Throughout February 2021, lagged estimates are about 2% below the positively biased convolutional ratios.

As primary incidence rises, we expect 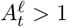, contributing positive bias relative to the well-specified ratio (Heuristic #1). Correspondingly, when hospitalizations surge due to the Delta variant in August 2021, the lagged ratio is less negatively biased. Lastly, recall we expect 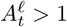 *after* a fall in primary incidence (Heuristic #3). This accounts for the lagged ratio having higher bias in April 2022, when hospitalizations level out from the Omicron surge. There, the lagged HFR (and presumably also the misspecification factor 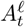) hits its maximum value.

We performed several robustness checks to assess the stability of these findings. Section S3 compares the ratio HFRs to multiple external estimates of the ground truth (Figure S4). It also studies the effect of using finalized count data (Figure S5) and different hyperparameter choices (Figures S6-8). Finally, it examines the ratio estimators’ behavior on six US states (Figure S9). By and large, the ratio estimators yield roughly the same type of bias throughout.

### 3.2 Simulated data

We further evaluate the ratio estimators in a variety of simulation settings. Keeping the primary incidence *X*_*t*_ as hospitalizations reported to the HHS, we simulate deaths based on the convolutional model (3) without noise. That is, we generate

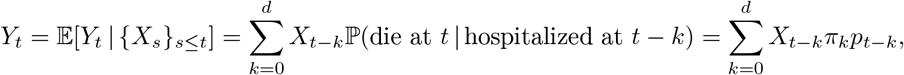

The simulations explore three different underlying HFR curves and two delay distributions. The delay distribution *π* is a discrete gamma with standard deviation 90% of its mean. We consider means of 12 and 24 to compare short and long distributions. For the HFRs *p*_*t*_, we first use the external estimates given by NHCS In addition, we mimic the opposite trend by inverting and rescaling this curve. We do so with the following formula, preserving the minimum and maximum HFRs:

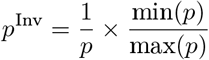

The third HFR setting is a stationary *p* = 10% over all time. This case elucidates the quantity 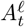 that drives the lagged ratio’s bias. This value is 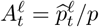, the lagged ratio itself scaled by a constant. To see this, recall that the oracle convolutional ratio is unbiased under stationarity, so Proposition 2 simplifies to 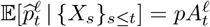 for the lagged ratio. Furthermore, 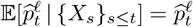 in this noiseless simulation, 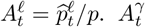so also is 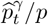 for the well-specified convolutional ratio, since the estimator is unbiased and 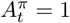 by definition.

To estimate the severity rates, the convolutional ratio is well-specified with *γ* = *π*. For the lagged ratio, we choose the lag 𝓁 by maximizing the cross-correlation between hospitalizations and deaths. In both cases, we do not smooth the data in any capacity. (The same is true for Figures 1 and 2.)

Figure 4 shows the results across the six settings in total. As expected, HFR estimates are significantly more biased when the underlying delay distribution is longer. This bias is most pronounced with the lagged ratio. For example, even when the true HFR is a constant 10%, the lagged ratio estimates hit 13% under the light-tailed delay, and 15% under the heavy-tailed one. In the NCHS HFR setting, it spikes (as we have seen before) as hospitalizations level out in spring 2022, reaching over 20% and 25% under the light- and heavy-tailed delays, respectively. Note the convolutional ratio does not share these dramatic oscillations. By and large, it tracks the general shape of the true curve, albeit at a delay. Its overall favorable performance rests on the fact that we have chosen in this simulation to provide it with the benefit of the true delay distribution (no misspecification).

The results in Section 2.3 explain the wide gap in performance between these estimators. As anticipated, the lagged ratio is higher than the oracle convolutional ratio when 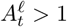, and lower when 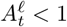. Comparing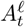 to the estimated HFR curves, the bias moves very similarly. For example, during the Delta and Omicron waves, rapid rises in hospitalizations produced high values of 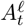. This accounts for the spikes in August 2021 and January 2022 (Heuristic #1). Heuristic #2 dictates the lagged estimator should have lower bias than the well-specified ratio as primary events fall. We observe this in Delta (September 2021) and Omicron (Februrary 2022). Lastly, when hospitalizations level out from the Omicron surge, 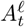 spikes to 1.2 and 1.5 for the short and long distributions. This explains the positive bias in spring 2022, per Heuristic #3.

The bias (7) rescales the oracle bias and adds a misspecification term. Studying Figure 4, we observe the misspecification term tends to dominate when 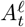 strays away from 1. To understand this, consider periods in which the oracle bias is negative. As introduced in Section 2.3, the oracle and misspecification terms are at odds with each other when this is the case. Invariably, the lagged ratio moves in the direction of the misspecification term 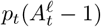. In the NHCS HFR setting, for example, the lagged estimates spike with 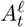 in August 2021. In the inverted setting, the lagged bias tracks the down-up-down motion of 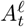 during the first five months of 2021. That the misspecification term wins out in these conflicting settings indicates it comprises a disproportionate amount of the bias. Indeed, the oracle bias is low enough that multiplicative rescaling must not have a large effect.

Section S4 of the Supplement repeats these analyses for CFR, using case data in lieu of hospitalizations (Figures S10-12). It shows the same qualitative behavior, with the well-specified convolutional ratio remaining decently calibrated and the lagged ratio displaying inaccurate, erratic swings. This indicates that the underlying bias mechanisms generalize directly to CFR and other severity rates.

## 4 Discussion

Our analyses and experiments illustrate that practitioners should take caution when using standard ratio estimators for time-varying severity rates. They exhibit nontrivial bias as severity rates change, particularly the popular lagged ratio. If a major purpose of such estimators is to inform stakeholders of changing risks in real time, then such bias indicates they may fail to do so in a reliable manner. While our main analyses focus on HFR, we additionally replicate the simulated experiments for CFR, confirming that ratio estimates of other severity rates share the same bias patterns.

Analyzing the bias, as we have done in Propositions 1 and 2, allows us to form real-time heuristics about what to expect in practice. For example, based solely on the primary incidence curve, we generally expect the lagged ratio to make the following errors:

- unreasonably high severity estimates when primary incidence is rising quickly (Heuristic #1);
- rapid declines when primary incidence is falling quickly (Heuristic #2);
- unexpected surges when primary incidence has leveled out after falling (Heuristic #3).

Practitioners may be able to adjust their reactions accordingly. For example, if the lagged CFR spikes shortly after hospitalizations have declined and reach a stable low point, then a savvy epidemiologist can temper their alarm with the knowledge it may well be spurious.

While the lagged ratio seems ubiquitous in practice, the convolutional ratio (when a reasonable estimate of the delay distribution can be formed) can be better behaved and should probably be favored. While it is still subject to bias, this tends to be of a smaller magnitude.

Going beyond, there is still room to improve upon the backward-looking convolutional ratio. A promising forward-looking severity estimator was proposed by Qu et al. ^(41)^. This method obtains all historical severity rates *p*_*t*_ at once, by minimizing

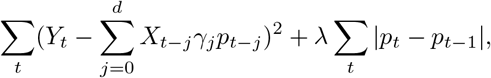

where *λ* ≥ 0 is a parameter that controls the level of regularization. The regularizer above is a total variation penalty, also called the fused lasso; it produces a piecewise constant fit of the severity rates. Unlike the convolutional ratio, this method models the relationship between events without assuming severity rates are locally stationary. Therefore, it may be a less biased alternative. Since the severity rates are defined implicitly via optimization, their bias is analytically intractable and must be assessed empirically.

Unfortunately, while this method was introduced as a real-time tool, it struggles with instability at the most recent timesteps. Improving its capabilities for real-time estimation, and extending it to fit smoother severity rates using trend filtering penalties (see, e.g., ^42^), are interesting directions for future work. Jahja et al. ^(36)^ applied trend filtering to a similar deconvolution problem, reconstructing latent infections from case reports. Their insights on tail regularization may be useful to stabilize real-time severity estimates.

Severity rates may be biased in ways beyond the statistical bias our work focuses on. In Section 2.4, we mentioned that HFR estimation from line-lists can be subject to “survivorship bias”: the failure to account for deaths occurring outside the hospital^(43)^. Under-reporting is another central challenge, particularly for CFR. Not all infections are reported, reporting rates change across time, and severe cases are more likely to be reported than mild cases^(44)^. Reich et al. ^(22)^ proposed an estimator for a time-invariant *relative* CFR—the ratio of CFRs between groups—which learns latent reporting rates via the EM algorithm. Angelopoulos et al. ^(45)^ applied this in the context of COVID-19. Their work also identifies other sources of bias, like differences in case definition and testing eligibility. Finally, examining how hyperparameter tuning strategies affect the bias poses an opportunity for future work.

As discussed in Section 2.1, severity rates may be understood in connection with reproduction numbers. This connection extends to their bias as well. For example, we demonstrated that the convolutional ratio is unbiased if the severity rate and delay distribution in the *d* time points before *t* are stationary. In a similar vein, Fraser ^(31)^ noted that instantaneous *R*_*t*_ is equal to case *R*_*t*_ if conditions remain unchanged. Future work along the lines of Eales and Riley ^(46)^ could apply our framework to *R*_*t*_ bias, examining the fidelity of instantaneous *R*_*t*_ as a proxy for case *R*_*t*_. Conversely, one could apply Bayesian perspectives on *R*_*t*_ to the context of severity rates.

## Data Availability

All data produced are available online at https://github.com/jeremy-goldwasser/Severity-Bias.

## Acknowledgements and Financial Disclosure

We would like to thank members of the Delphi research group for helpful feedback. This study was funded by two grants from the Centers for Disease Control and Prevention (CDC): “The Delphi Center for Outbreak Analytics and Disease Modeling in Public Health Response” (no. NU38FT00005), and “Digital Public Health Surveillance for the 21st Century” (no. 75D30123C15907). Funders were not involved in this study in any capacity. Ryan Tibshirani received both grants, and used them to fund Jeremy Goldwasser.

## Supporting Information Legends

**Figure S1**. Toy examples demonstrating biased severity-rate estimators under different delay structures and primary-incidence patterns. Panels show: (a) all deaths after 𝓁 days for 𝓁 = 14 and 28; (b) changing primary incidence and bias of lagged vs. convolutional ratios; (c) sinusoidal severity rates; (d) step-function severity shifts.

**Figure S2**. Severity rates where the true delay distribution is time-varying. Dashed lines use true nonstationary delay distributions; dotted lines use constant distributions and lags.

**Figure S3**. Comparison of lagged ratio estimates based on JHU versus NCHS death data.

**Figure S4**. External HFR estimates from: retrospective NCHS-based ratios and variant-mixed HFRs, compared to NHCS estimates.

**Figure S5**. Comparison of estimates based on real-time versus finalized hospitalization and death counts.

**Figure S6**. Sensitivity of lagged and convolutional ratios to post-smoothing window lengths.

**Figure S7**. Lagged HFR estimates across different lag parameters (2–5 weeks).

**Figure S8**. Convolutional ratio estimates under different delay distributions, varying mean and standard deviation.

**Figure S9**. State-level HFR estimates comparing real-time convolutional ratios, lagged ratios, and retrospective NCHS-based estimates for six large U.S. states.

**Figure S10**. CFR analyses mirroring Figure 1: effects of changing severity, delay distributions, and primary incidence on the well-specified convolutional ratio.

**Figure S11**. CFR misspecification effects under light-tailed, heavy-tailed, and lagged delay distributions. **Figure S12**. CFR ratio estimates under various simulation settings, paralleling Figure 4: comparison of oracle convolutional and lagged ratios across multiple severity scenarios.

## Footnotes

1 Of course, conditions in the UK may be quite different from the US. However, we rely on the UK study because it provides the most comprehensive information on COVID-19 hospitalization-to-death delay distributions.

