## Supplement for "Challenges in Estimating Time-Varying Epidemic Severity Rates from Aggregate Data"

### Supplemental Information for *Challenges in Estimating Time-Varying Epidemic Severity Rates from Aggregate Data*

#### S1 Proofs

The assumption of stationary delay distribution is not necessary for either bias expression. In the proofs in Sections S1.1 and S1.2, the delay distributions $\pi$ may simply be replaced with $\pi^{(t)}$.

##### S1.1 Proof of Proposition 1

Proposition 1 establishes the bias of the well-specified convolutional ratio. Here and henceforth, we abbreviate $\mathbb{E}_t[\cdot] = \mathbb{E}[\cdot \mid \{X_s\}_{s \le t}]$. Observe that

$$\begin{aligned}
\mathrm{Bias}(\widehat{p}_t^{\pi}) &= \frac{\mathbb{E}_t[Y_t]}{\sum_{k=0}^d X_{t-k}\pi_k} - p_t \\
&= \frac{\sum_{k=0}^d X_{t-k}\pi_k p_{t-k}}{\sum_{k=0}^d X_{t-k}\pi_k} - \frac{p_t \sum_{k=0}^d X_{t-k}\pi_k}{\sum_{k=0}^d X_{t-k}\pi_k} \\
&= \sum_{k=0}^d \frac{X_{t-k}\pi_k}{\sum_{j=0}^d X_{t-j}\pi_j}(p_{t-k} - p_t).
\end{aligned}$$

#### S1.2 Proof of Proposition 2

Proposition 2 establishes the bias of the misspecified convolutional ratio, the lagged ratio being a special case. Observe that

$$\begin{aligned}
\text{Bias}(\widehat{p}_t^{\gamma}) &= \frac{\mathbb{E}_t[Y_t]}{\sum_{k=0}^{d} X_{t-k}\gamma_k} - p_t \\
&= \frac{\sum_{k=0}^{d} X_{t-k}\pi_k p_{t-k}}{\sum_{k=0}^{d} X_{t-k}\gamma_k} - \frac{\sum_{k=0}^{d} X_{t-k}\gamma_k p_t}{\sum_{k=0}^{d} X_{t-k}\gamma_k} \\
&= \sum_{k=0}^{d} \frac{X_{t-k}}{\sum_{j=0}^{d} X_{t-j}\gamma_j}(\pi_k p_{t-k} - \gamma_k p_t) \\
&= \sum_{k=0}^{d} \frac{X_{t-k}}{\sum_{j=0}^{d} X_{t-j}\gamma_j}(\pi_k p_{t-k} - (\pi_k + (\gamma_k - \pi_k))p_t) \\
&= \frac{\sum_{j=0}^{d} X_{t-j}\pi_j}{\sum_{j=0}^{d} X_{t-j}\gamma_j} \sum_{k=0}^{d} \frac{X_{t-k}\pi_k}{\sum_{j=0}^{d} X_{t-j}\pi_j}(p_{t-k} - p_t) - p_t \sum_{k=0}^{d} \frac{X_{t-k}}{\sum_{j=0}^{d} X_{t-j}\gamma_j}(\gamma_k - \pi_k) \\
&= \frac{\sum_{j=0}^{d} X_{t-j}\pi_j}{\sum_{j=0}^{d} X_{t-j}\gamma_j} \text{Bias}(\widehat{p}_t^{\pi}) + p_t \left( \frac{\sum_{k=0}^{d} X_{t-k}\pi_k}{\sum_{j=0}^{d} X_{t-j}\gamma_j} - 1 \right).
\end{aligned}$$

### S2 Additional analysis and data sources

#### S2.1 Further analysis of bias

We first present examples that further explain the bias for the well-specified convolutional ratio. These examples are considerably more contrived that the ones in Section 2.3. Nonetheless, their bias can be simplified to simple analytic formulas, isolating the three contributing factors.

To elucidate the relationship between changing severity rates and bias, let us consider the case where all secondary events occur after precisely $\ell$ time points. The well-specified convolutional and lagged ratio estimators coincide: $\widehat{p}_t^{\gamma} = \widehat{p}_t^{\ell} = p_{t-\ell}$. The bias in this setting is simply the change in the true severity rate, $p_{t-\ell} - p_t$, and the ratio estimator is unbiased only if the severity rate is stationary. Otherwise the ratio will be 20% too low, for example, if the true severity rate was 20% lower $\ell$ time steps ago.

Figure S1a displays the results on the NHCS HFRs. In general, severity rates will be less similar to the present value $p_t$ as we go further back in time. The bias $p_{t-\ell} - p_t$ tends to be larger when $\ell = 28$ versus $\ell = 14$. This supports the overarching idea that estimates with heavier-tailed delay distributions tend to have more bias.

Now to elucidate the relationship between primary incidence and bias, let us consider a delay distribution $\pi$ which places half its mass at lag 0, and the other half at lag $q$. Then the well-specified bias has magnitude:

$$\left|\text{Bias}(\widehat{p}_t^{\pi})\right| = \frac{\frac{1}{2}\left|X_t(p_t - p_t) + X_{t-q}(p_{t-q} - p_t)\right|}{\frac{1}{2}(X_t + X_{t-q})} = \frac{|p_{t-q} - p_t|}{1 + X_t/X_{t-q}}.$$

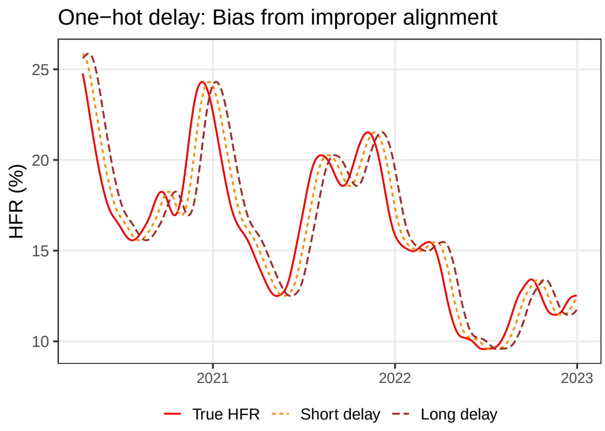


(a) All deaths after $\ell$ days. HFR ratios equal; plotting delays of $\ell = 14$ and 28 days.

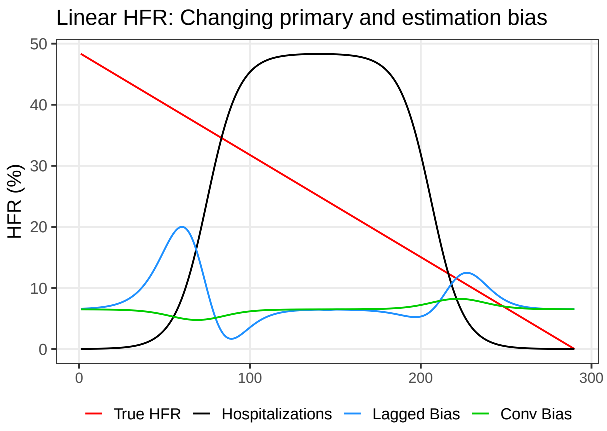


(b) Changing primary incidence. Bias of lagged and convolutional ratios.

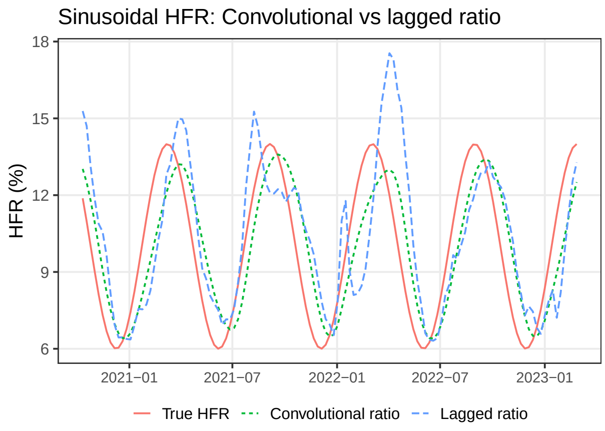


(c) Sinusoidal severity rate and ratio estimates.

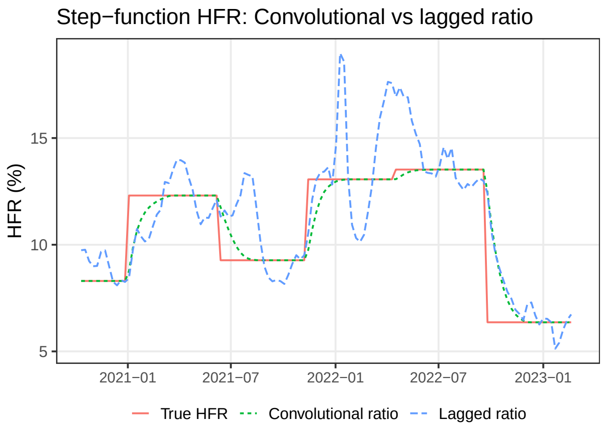


(d) Severity rate shifts via step functions.

Figure S1: Toy examples demonstrating biased severity-rate estimators under different delay structures and primary-incidence patterns.

In other words, the absolute bias is monotonically decreasing in $X_t/X_{t-q}$, the proportion change in primary incidence. Rising primary incidence ($X_t/X_{t-q} > 1$) yields less bias, while falling levels yield more.

Figure S1b displays this setting with $q = 10$. Rising hospitalizations are defined as $X = \sigma(s) * 9000 + 1000$, where $\sigma$ is the sigmoid function and $s$ takes 300 evenly spaced steps from -9 to 7. These quantities are subsequently reflected to express a decline. The true HFRs fall from 0.5 to 0 over the same number of even steps. Accordingly, the absolute bias of the convolutional ratio is $c_q \frac{X_{t-q}}{X_{t-q}+X_t}$, where $c_q \approx 0.0167$. Shown in red, it dips as hospitalizations rise, and rises as they fall.

The figure also plots with lagged ratio with $\ell = \frac{d}{2}$, the mean of the delay distribution. When daily hospitalizations are close to constant, the two estimators converge towards the same ratio. During periods of change, however, the lagged estimator has different bias. It first moves upwards — the opposite direction as the convolutional bias — with far greater magnitude. This can be explained by the ratio $A_t^\ell = \frac{X_{t-2\ell}+X_t}{2X_{t-\ell}}$ from Proposition 2. As hospitalizations begin to steeply rise, $X_{t-2\ell}$ and $X_{t-\ell}$ are similar, but $X_t > X_{t-\ell}$. Hence, $A_t^\ell > 1$, contributing positive bias to both the oracle and misspecification terms. As hospitalizations level out near the top, $A_t^\ell < 1$, hence the bias falling lower. The opposite pattern occurs as hospitalizations fall.

We consider two additional settings with contrived severity rates, beyond the linear change in Figure S1b. Figures S1c and S1d generate deaths with sinusoidal and piecewise constant HFRs, respectively. They use true US hospitalization counts throughout over two years of the COVID-19 pandemic. Deaths are simulated noiselessly to highlight the estimators' bias. The delay distribution is the same as in the simulated experiments in the main text: A gamma distribution whose mean maximizes the correlation between hospitalization counts from HHS and death counts from JHU.

Figure S1c reveals the ratio estimators have varying capacity for bias when the true severity rate is sinusoidal. In general, the ratios trail behind the true values some number of days. The convolutional ratio mimics the general sinusoidal shape, though its high and low values are about 1% less stark than the true values. Intuitively, the ratio smooths over the trailing history, so it is incapable of precisely identifying peak and trough values. The lagged ratio, as explained in the main text, depends heavily on the primary incidence curve and its relation to the delay distribution. Consistent with our analysis in Section 3.2, this leads to high values of $A_t^\ell$ inflating HFR estimates in Spring 2022. We see the highest lagged HFR reaching 17%, while the true severity rate turns over around 14%.

Figure S1d tracks the ratios as the true severity rate jumps up and down by varying degrees. The well-specified convolutional ratio gradually reaches each new plateau and stabilizes. The time for it to adjust depends on the magnitude of the shift. This is consistent with our analysis in Section 2.2, which showed that larger changes in severity rate correspond to more bias.

The lagged ratio is more heavily dependent on the primary incidence curve, acting through $A_t^\ell$. Proposition 2 shows this term alters the well-specified bias via a multiplicative correction and, more significantly, an additive adjustment. How $A_t^\ell$ produces the lagged ratio's high positive, negative, and positive bias with a constant severity rate in early 2022 is explained precisely in Section 3.2. Another example of biased divergence occurs in the Delta wave of summer 2021, in which rising primary incidence triggers a rise in $A_t^\ell$ and thus $\widehat{p}^\ell$. However, when primary incidence is roughly constant, the lagged and convolutional ratios coincide. We see this in the latter half of 2022, when hospitalization counts were low.

We additionally evaluated these methods when the true delay distribution is time-varying. Our analyses focus on the bias at a single point in time, so their messages should transfer translate to the time-varying setting. To generate realistic delay distributions, we identified the major variant periods using data described in Section S3.1. Within these ranges, we computed the correlation-maximizing lag between HHS hospitalizations and JHU deaths. These lags ranged by a surprising margin: only a handful of days for the Delta wave, and over 30 during Omicron. We then computed the expected hospitalization-to-death delay by mixing these lags with the proportions of each variant in circulation. (This is akin to how we alternatively estimated HFRs in the aforementioned Section.) Finally, we parameterized the delay distributions based on these mean delays and their corresponding standard deviations.

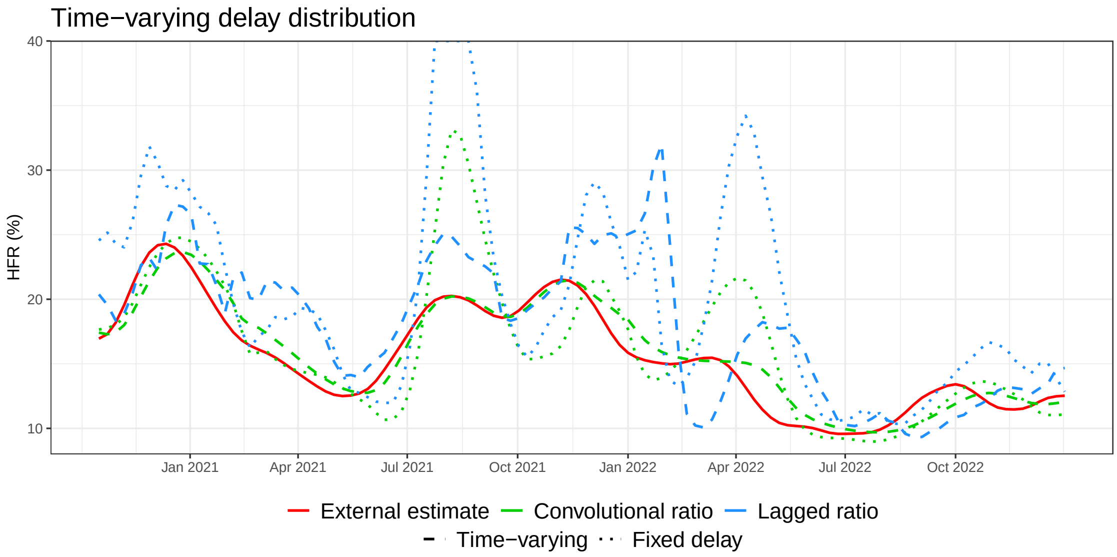


Figure S2: Severity rates where the true delay distribution is time-varying. Dashed lines compute ratios with the true nonstationary delay distributions and their means as the lags. Dotted lines use the same constant distribution and lag – their means over all time.

We simulated deaths without noise from these delay distributions and the NCHS-based HFRs.

On this simulated data, we computed the convolutional and lagged ratios in two ways. Firstly, we let them use the true time-varying delay distributions and lags. Under this formulation, the convolutional ratio is well-specified; the lagged ratio is not, as it still reduces the distribution to a point mass, but the mass is at least selected in the proper location. The second strategy ignored the time-varying nature of the true model. It instead used stationary quantities: the mean hospitalization-to-death distribution and lag (18 days) over all time. This stationarity induces misspecification bias into both ratio estimators.

The behavior of the biases in Figure S2 is consistent with our other analyses. The well-specified convolutional ratio is close to unbiased during stretches for which the delay distribution had low mean. During Omicron, when deaths were reported at a significant delay, it tracks the ground truth HFR at a much longer offset. This observation in the time-varying case is identical to our analysis that the well-specified convolutional ratio is more biased under heavier-tailed delay distributions (see, for example, Figure 1b).

The corresponding lagged ratio is also more biased during Omicron than Delta in this experiment. The light-tailed delay distribution indicates the probability mass is more condensed around the lag, so $A_t^\ell$ is not significantly greater than 1 as hospitalizations rise in Delta. However, as Omicron surged around the start of 2022, the lag is over 30 days. Hospitalizations were much lower at this offset, so $A_t^\ell$ has the capacity to get very large, and thus produce a high amount of bias. The bias is greatest around the peak in late January, when the lagged ratio is more than double the true HFR of 15%. Interestingly, the positive bias that spring is less pronounced than in previous experiments. This is because the longer lag here captures more distant hospitalizations, which brings down $A_t^\ell$ somewhat.

The misspecified ratios with stationary parameters exhibit more bias. In the Delta wave, where the delay distribution and lag were too long, both estimators were much too high. They placed too much emphasis on low counts as the surge was beginning, resulting in significant positive bias. This was particularly pronounced for the lagged ratio, which was positively biased even under the accurate lag. It peaked off the figure at 60%, triple the true HFR.

During Omicron, the ratios' delay distributions and lags were too short. This led to *less* bias than the appropriately-specified ratios during the surge and decline, since these parameters accurately reflected current conditions. However, it produced high positive bias for both estimators after the surge leveled out in April 2022. As in the main text, the lagged ratio is extremely biased, since its denominator entirely fails to reflect the bygone surge. It reaches over 30% as the true HFR falls towards 10%.

#### S2.2 Retrospective deaths

JHU aggregated daily deaths in real time, aligned by the date they were reported. In contrast, the National Center for Health Statistics (NCHS) provided weekly totals of deaths aligned by occurrence, which were not available in real time. Intuitively, the delay which relates hospitalization to death occurrence should have a lighter tail in comparison to that relating hospitalization to death report. Therefore, since that heavier-tailed delays generally introduce greater bias, we would expect the ratio estimates computed from JHU deaths to have greater bias than those from NCHS deaths. Figure S3 shows that this is indeed the case. NCHS data leads to lagged HFR estimates with substantially less bias. However, this resource was only available in retrospect, so these "real-time" estimates could not actually be produced until long after the dates in question.

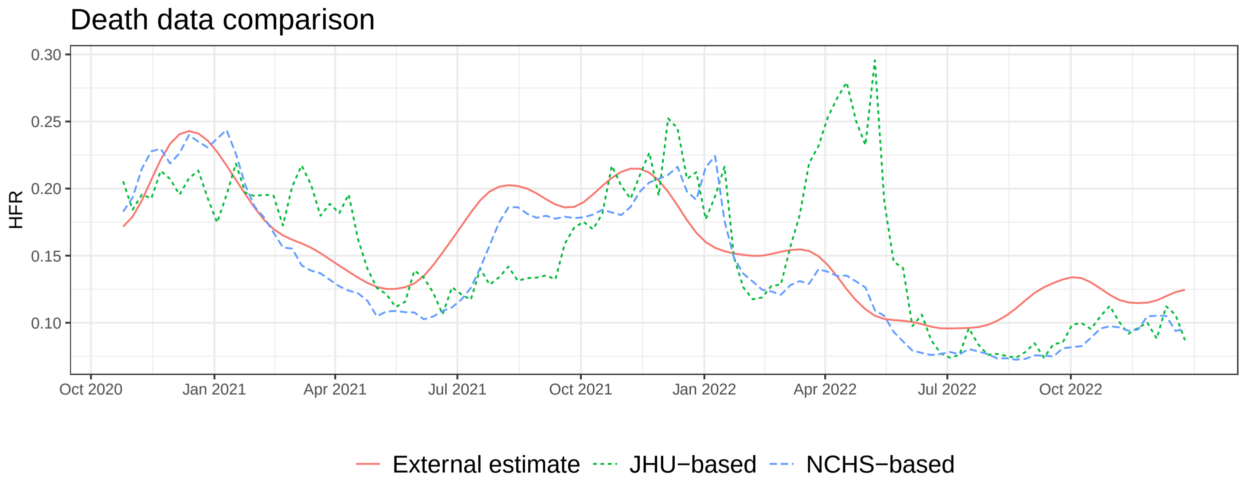


Figure S3: Comparing lagged ratios based on data from JHU versus NCHS.

### S3 Robustness checks

#### S3.1 Alternative external estimates of HFR

We consider two alternative approaches to approximate the true HFR curve, over the COVID-19 pandemic. The first is simply to use the lagged ratio based on NCHS data. As discussed above, this benefits from a lighter-tailed delay distribution than JHU data. Further, as we able to use data after $t$ to estimate $p_t$, we use the retrospective severity estimator introduced in Overton et al., 2022:

$$\widehat{p}_t = \sum_{k=0}^{d} \frac{Y_{t+k}\widehat{\pi}_k^{(t)}}{\sum_{j=0}^{d} X_{t+k-j}\widehat{\pi}_j^{(t+k-j)}}. \tag{1}$$

The second approach we consider is to compute a single HFR by dividing total deaths by total hospitalizations in each major variant period, and then create a smooth curve by mixing these per-variant HFRs by estimates of the proportion of variants in circulation, obtained from CoVariants.org. We only consider the four largest variants: the original strain, Alpha, Delta, and Omicron. However, because Omicron began with an enormous surge that quickly subsided, we split it into early and late periods.

Figure S4 displays these two alternative HFR estimates, alongside the curve obtained from NHCS. They have nontrivial differences in magnitude, but reassuringly, the three curves move more or less in conjunction. The retrospective NCHS ratio is still subject to statistical bias similar to equation (7); the variant-based HFR curve is flatter, as it does not account for other sources of potential variability in the underlying severity rate.

#### S3.2 Real-time versus finalized data

Recall, the results in Section 3.1 use hospitalization and death counts available in real time. To investigate the sensitivity of our findings, we recompute the lagged and convolutional ratios, this time using finalized counts. Figure S5 shows the real-time and finalized estimates

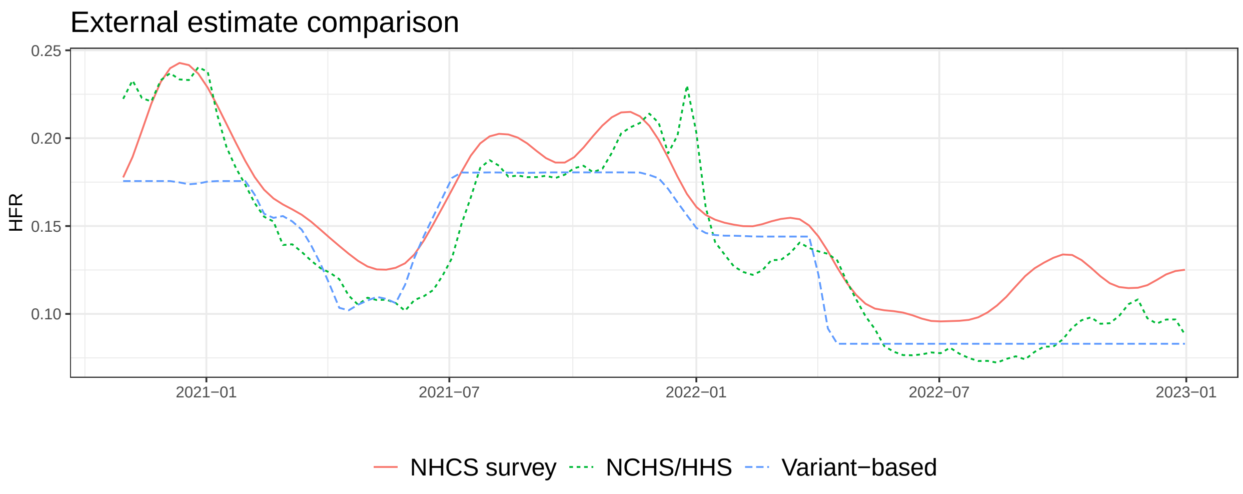


Figure S4: Comparing methods that use external data to retrospectively approximate HFR.

track one another very closely. Therefore, the observed bias in Figure 3 cannot be attributed to real-time reporting quirks.

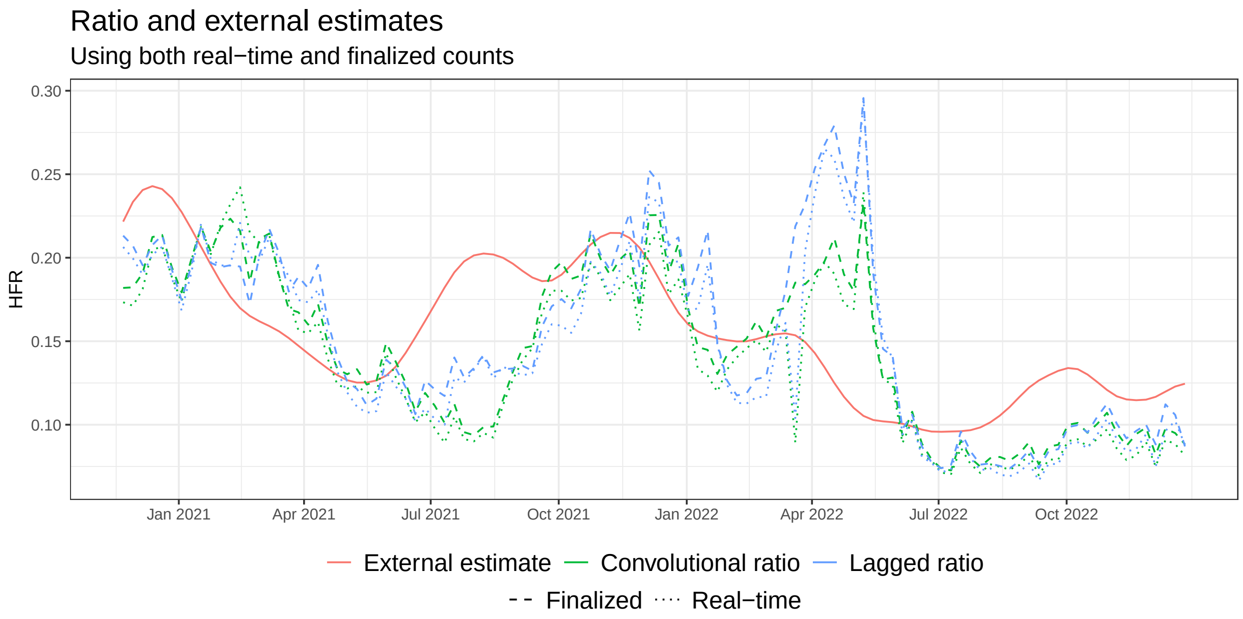


Figure S5: Comparing estimates based on real-time versus finalized counts.

#### S3.3 Hyperparameters

We evaluate the robustness of our findings against choices of hyperparameters. First, we analyze smoothed versions of the ratio estimators, where we smooth the numerator and denominator separately:

$$\widehat{p}_t^{\ell,w} = \frac{\sum_{s=t-w+1}^{t} Y_s}{\sum_{s=t-w+1}^{t} X_{s-\ell}},$$

$$\widehat{p}_t^{\gamma,w} = \frac{\sum_{s=t-w+1}^{t} Y_s}{\sum_{s=t-w+1}^{t} \sum_{k=0}^{d} X_{s-\ell-k}\gamma_k}.$$

Figure S6 shows the results for varying window lengths $w > 0$. The results are very similar, indicating the bias does not disappear when smoothing over a longer history.

We next examine the time-to-death hyperparameters: The lag for the lagged ratio and delay distribution for the convolutional ratio. Figure S7 displays lagged HFR estimates where the lag $\ell$ ranges from 2 to 5 weeks. Unlike the window size, changing this parameter leads to notably different behavior. Some choices of lag are better than others; a 28-day lag, for example, falls appropriately in winter 2021, and rises less slowly during Delta. However, all are biased to varying degrees, most notably the huge spurious surge in spring 2022.

Figure S8 compares the performance of the convolutional ratio across different choices of delay $\gamma$; we kept the discrete gamma shape for each, but varied the mean and standard deviation. We investigated a standard deviation equal to 90% of the mean, and also a more

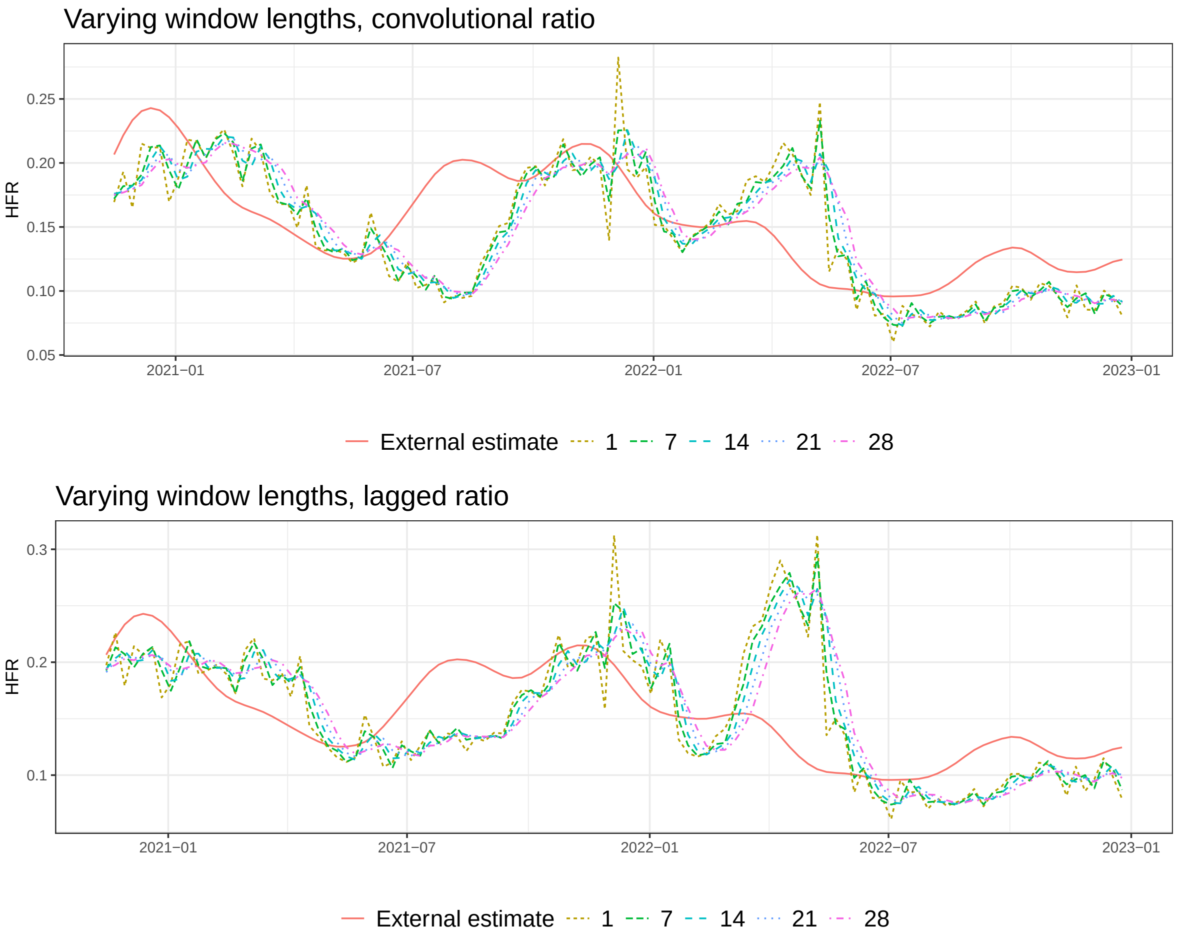


Figure S6: Comparing different choices of window length in post-smoothing.

compact delay with standard deviation equal to 70% of the mean. All resulting HFR estimates are significantly biased. Regardless of delay distribution, the ratios are negatively biased during the onset of Delta, and surge after the peak of Omicron. This indicates the bulk of the error is fundamental to the estimator, and cannot be attributed to model misspecification.

#### S3.4 State-level results

We repeat our analysis the six large US states, finding similar trends. The NHCS survey was conducted on a subset of hospitals meant to represent the US at large, so it cannot be used to accurately estimate the HFRs within each state. Instead, our external estimates of the state-level HFRs use the retrospective ratio discussed above (Equation 1), with NCHS deaths. Figure S9 compares this external HFR estimate to the real-time convolutional and lagged ratios. For each state, we select the lag for the real-time lagged ratio to maximize cross-correlation between hospitalizations and JHU deaths. We then use a discrete gamma distribution with mean equal to this lag, and standard deviation 90% of its mean, for the convolutional ratio.

Several states display similar biases to the US as whole (Figure 3). Estimates in California,

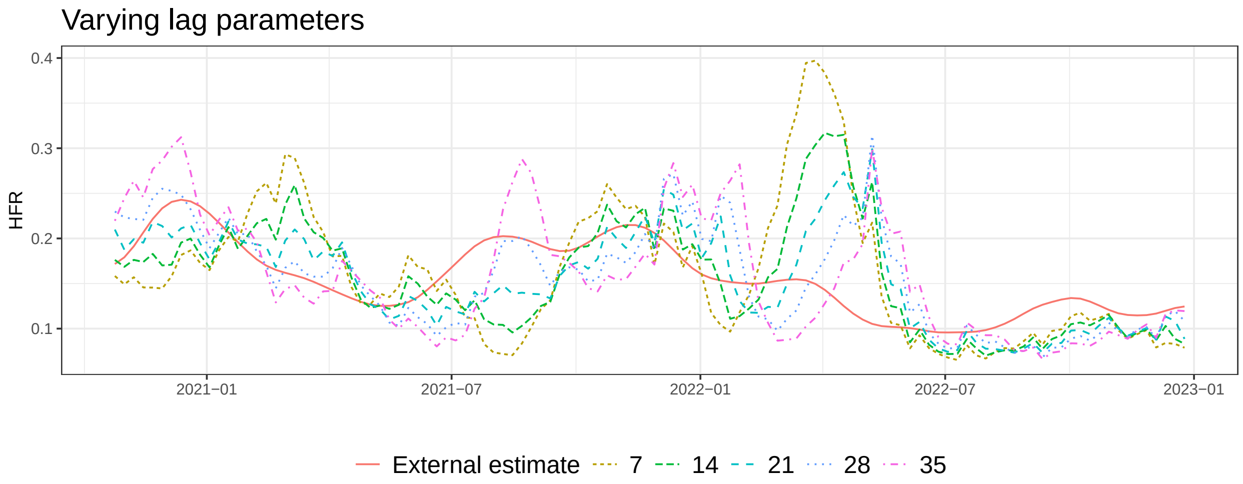


Figure S7: Comparing different choices of lag parameter in the lagged ratio.

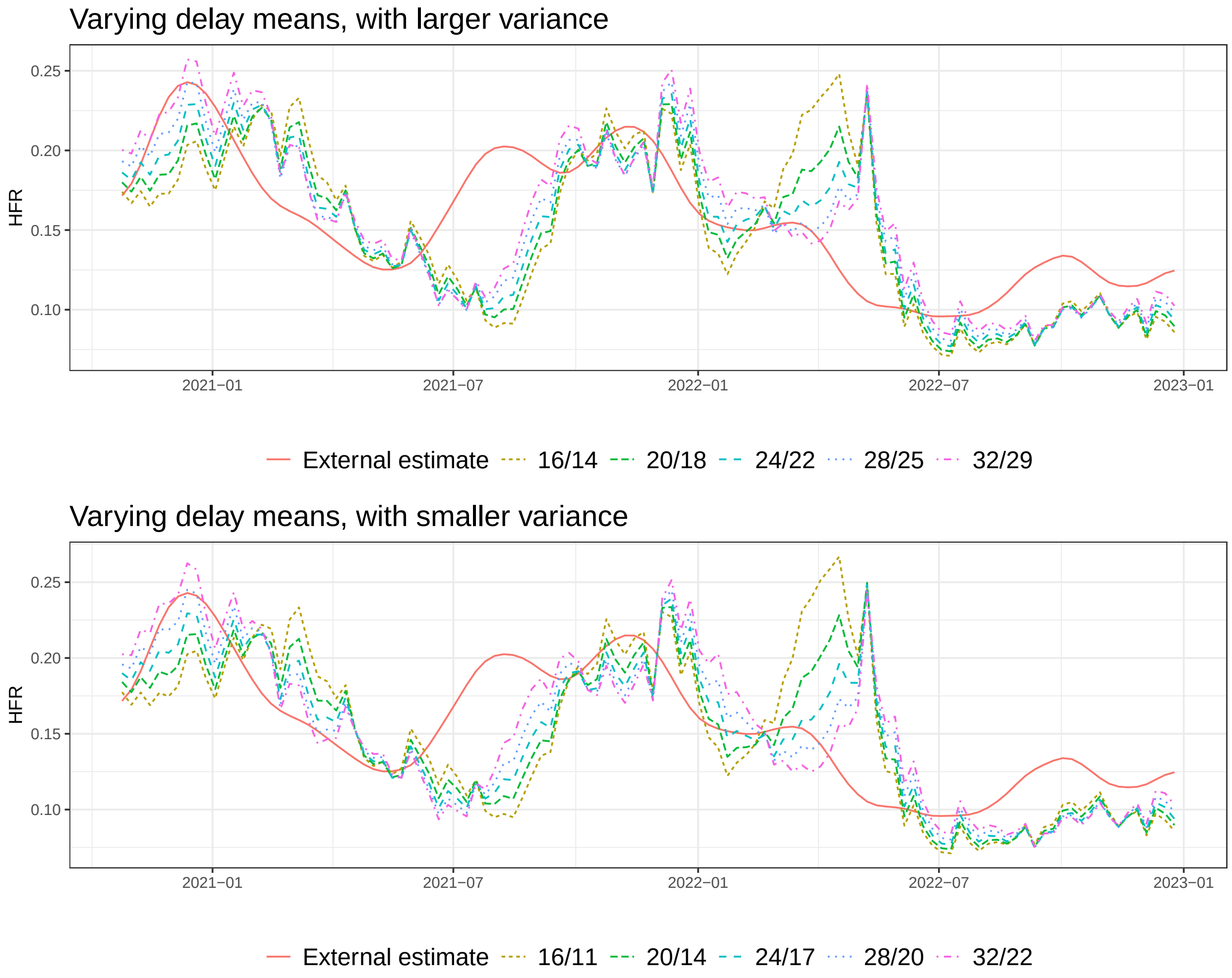


Figure S8: Comparing different choices of delay distributions in the convolutional ratio. The top panel shows gamma distributions whose standard deviation is 0.9 times the mean, versus 0.7 times the mean the bottom panel. The legend labels reflect the mean/standard deviation.

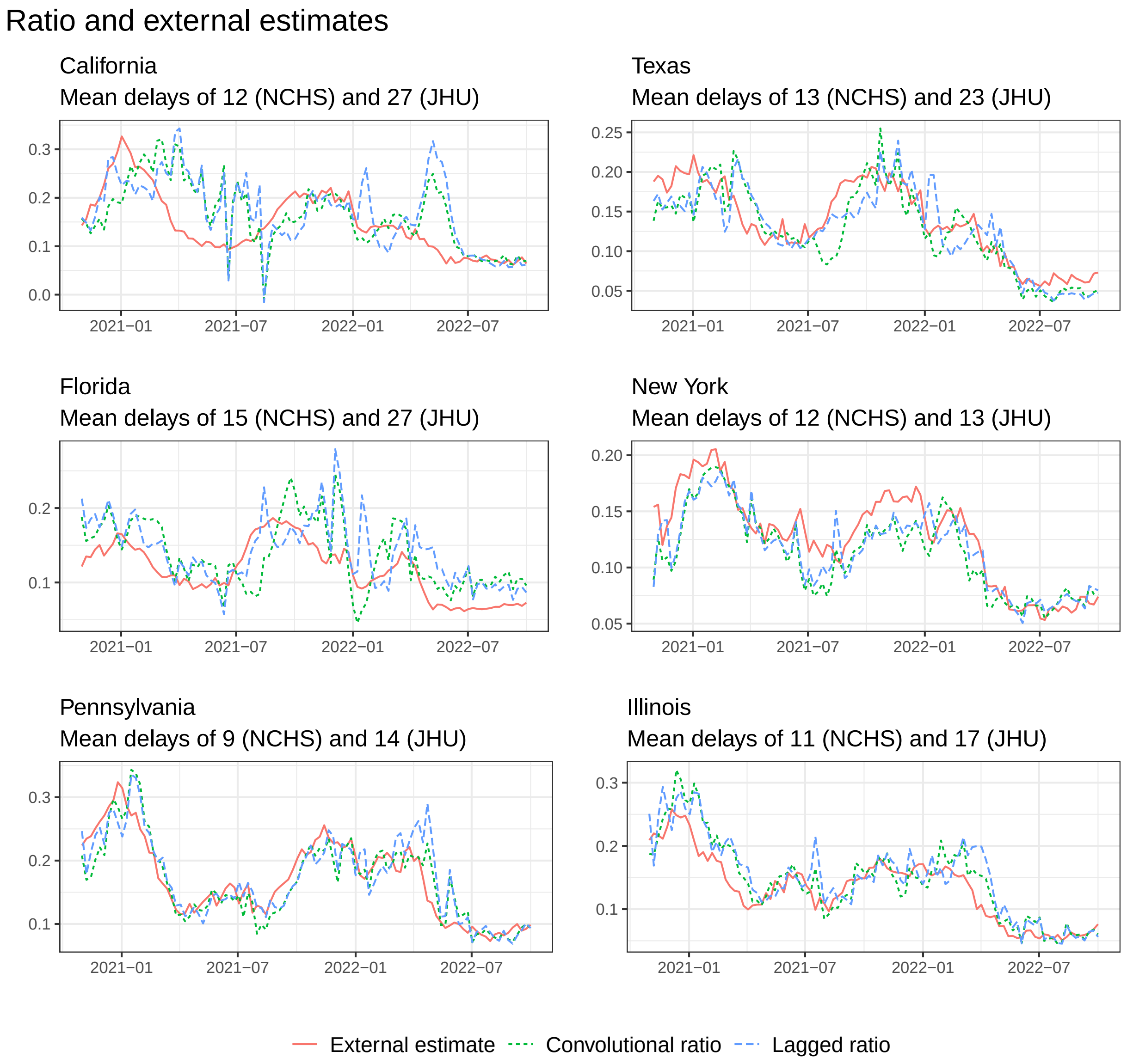


Figure S9: Comparing ratio estimates for individual US states.

Texas, and Florida are all slow to detect the uptick in HFR during Delta; they also spike during Omicron in California, and to a lesser extent Florida. Note these states are the ones with the largest cross-correlation optimal lags, an estimate of the average time-to-death. In contrast, New York, Pennsylvania, and Illinois have mean delays of at most 17. Their HFR estimates are still biased, but overall less so. This once again emphasizes the role of the delay in bias. The takeaway: fatality ratios are generally less trustworthy in states that take longer to report deaths.

### S4 CFR analysis

To illustrate the ubiquity of our analyses across all severity rates, we recreated our simulated experiments with CFR as opposed to HFR. As a recap, the case-fatality rate (CFR) measures the proportion of reported cases that ultimately die. It is often used as a proxy for the infection-fatality rate (IFR), since true infection counts are challenging to estimate, especially in real-time.

In this section, we recreate Figures 1, 2, and 4, using cases instead of hospitalizations. We restrict our focus to simulated deaths because unlike HFR, a strong external estimate for the true CFR is unavailable in real-time. Another reason for this restriction is that CFR is a problematic surrogate for IFR. Not all infected individuals show up as positive case reports, and the case ascertainment rate varies over time. Therefore, devising a schema to estimate IFR on COVID-19 data in real-time, and comparing it to notions of the "true" IFR and CFR is out of scope for this work.

The ground truth CFRs in these simulations follow the same NHCS-based curve in our prior experiments. To reflect CFRs, we rescaled this curve such that the average CFR was the overall COVID-19 CFR from 2020-2022. Deaths are simulated using the same delay distributions as in HFR. This is justified by our confirmation that the correlation-maximizing lag between US cases and deaths is the same as that between hospitalizations and deaths.

Figure S10 presents the CFR counterpart to Figure 1 in the main text. As with HFR, the same qualitative trends emerge: rapid changes in the underlying severity rate produce larger biases, longer delay distributions amplify these distortions, and variations in the primary incidence curve modulate both the direction and magnitude of bias. These parallels confirm that the mechanisms driving bias in the well-specified convolutional ratio are not specific to HFR but generalize directly to CFR.

Figure S11 shows the effects of delay-distribution misspecification for CFR. The same qualitative patterns observed for HFR persist: the light-tailed delay yields upward or downward deviations depending on incidence trends, the heavy-tailed delay has smaller opposing bias, and the lagged ratio remains the most volatile. These results confirm that the misspecification mechanisms outlined in Proposition 2 extend directly to CFR.

In fact, the bias is even larger than in the HFR case. The lagged ratio spikes to 2.5%, a staggering 150% above the true CFR of 1%. In contrast, lagged HFRs peaked at 25% while the true values were 15% The behavior of $A_t^\ell$ explains this discrepancy. It peaks around 2.5 for CFR, compared to 1.5 for HFR. The larger value for CFR is driven by cases climbing higher and falling steeper than hospitalizations, in relative terms.

Figure S12 shows the simulated results for CFR, paralleling Figure 4 from the main text.

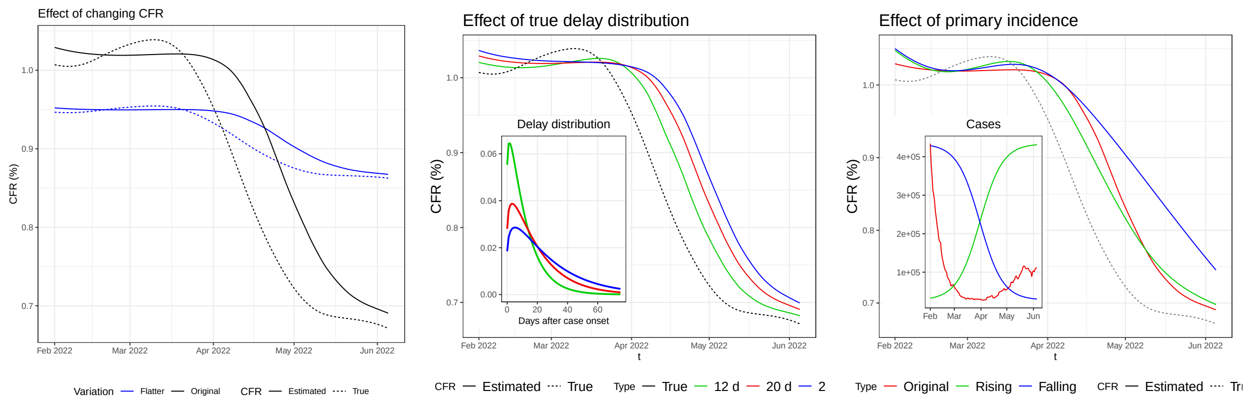


(a) (b) (c)

Figure S10: These figures illustrate the same qualitative trends as in Figure 1, now using CFR instead of HFR. The same three factors—changing severity, delay distribution, and primary incidence—produce similar effects on the bias of the well-specified convolutional ratio.

The qualitative behaviors mirror those observed for HFR, confirming that the same bias mechanisms apply to CFR-based analyses. The convolutional ratio continues to track the true curve relatively closely, while the lagged ratio exhibits exaggerated oscillations, especially under the longer delay. When incidence surges, the lagged CFR overshoots the truth; when incidence falls, it undershoots. These dynamics follow directly from the misspecification behavior of $A_t^\ell$ described in Proposition 2.

Even in the stationary-severity simulations, the lagged ratio remains heavily biased upward whenever $A_t^\ell > 1$. Its peaks for CFR roughly twice as large as those seen for HFR. As discussed for Figure S11, this difference is driven by the sharper swings in case incidence compared with hospitalizations. Meanwhile, the convolutional ratio's stability reflects its correct specification of the delay distribution—its advantage holding equally for CFR as for HFR.

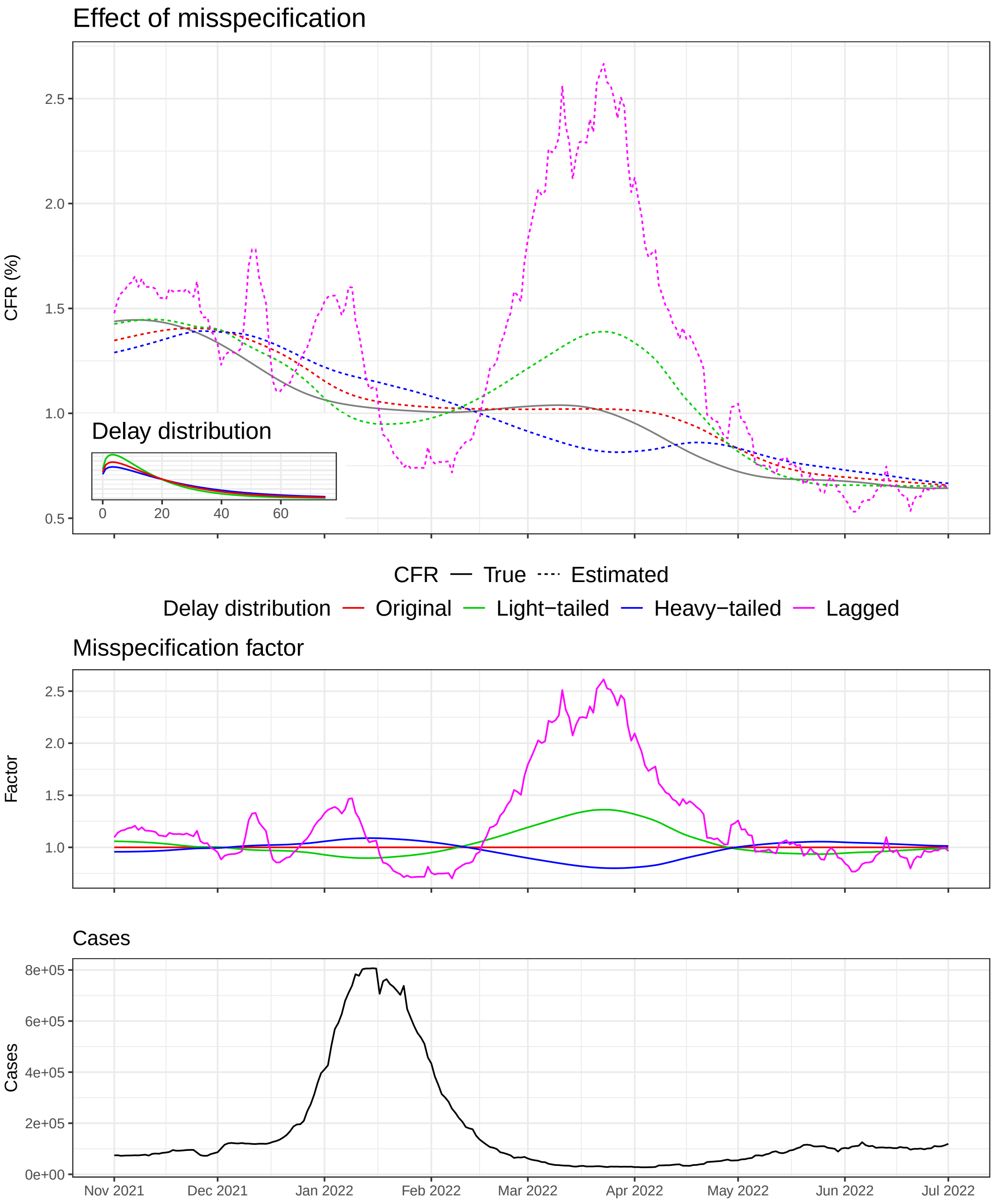


Figure S11: Analogous to Figure 2, this figure illustrates the effects of delay-distribution misspecification when using CFR instead of HFR. The same qualitative behavior is observed across light-tailed, heavy-tailed, and lagged specifications.

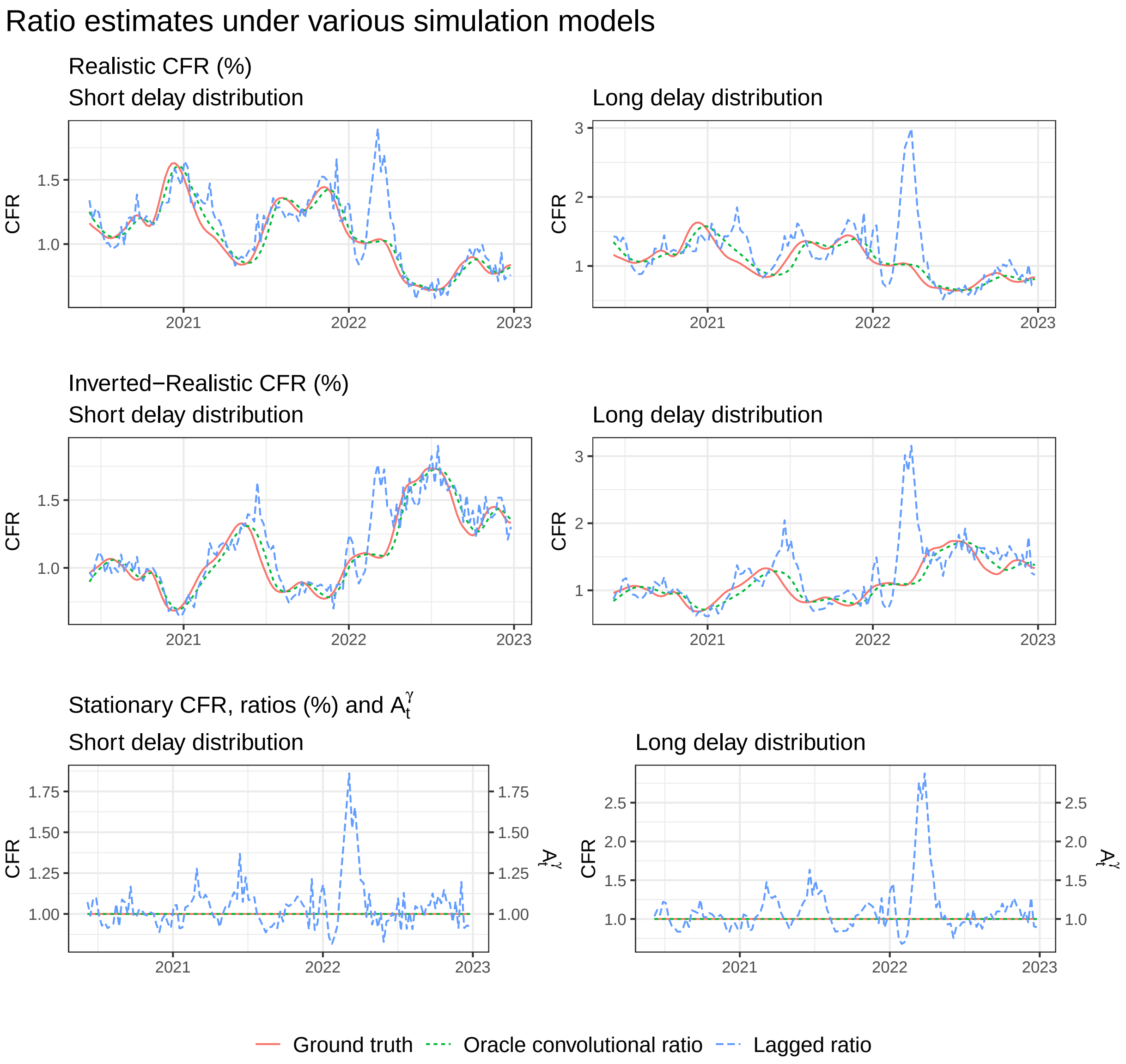


Figure S12: Ratio estimates under various simulation settings, using CFR instead of HFR. As in Figure 4, the convolutional ratio tracks the true severity curve closely, while the lagged ratio exhibits larger oscillations, particularly under the longer delay.
